# Comorbidity patterns and sex differences in late-onset Alzheimer’s disease using electronic records

**DOI:** 10.1101/2025.04.24.25326306

**Authors:** Yukari Katsuhara, Umair Khan, Zachary A. Miller, Isabel E. Allen, Tomiko T. Oskotsky, Marina Sirota, Alice S. Tang

## Abstract

In this retrospective study, we applied unsupervised learning techniques to electronic medical records from UCSF to characterize Alzheimer’s disease comorbidity patterns. Given the well-known female predominance in Alzheimer’s disease, we performed sex-stratified analyses to explore differences in comorbidity patterns based on sex. Findings were evaluated using an independent UC-Wide dataset. Among 8,363 patients in the UCSF dataset, we observed five data-driven comorbidity clusters related to cardiovascular conditions, gastrointestinal disorders, and frailty-related conditions such as pneumonia and pressure ulcers, as well as groups with higher and lower overall comorbidity burden. Sex-stratified analyses within clusters identified variation in comorbidity patterns, including circulatory diseases in males in Cluster 2 and bladder stones in females in Cluster 3. Key results demonstrate partial consistency in the UC-Wide dataset. Our study provides a structured characterization of comorbidity heterogeneity in AD and highlights sex-related differences, though findings should be interpreted as descriptive and hypothesis-generating.

**Highlights:**

- Among an EHR cluster analysis, Alzheimer’s disease patients stratify into five data-set derived comorbidity clusters.
- Clinical patterns and comorbidity burdens vary across patient subgroups.
- Sex-specific differences influence comorbidity profiles in selected patient clusters.
- Highly granular cluster configurations lack robust cross-dataset reproducibility.

## Introduction

Alzheimer’s disease (AD) is the most common cause of dementia. With rising prevalence and no curative treatments, AD represents a major public health crisis worldwide. Clinically, AD has traditionally been conceptualized as an amnestic disorder predominantly affecting memory in those 65 and older, but it can also manifest at an earlier age and with non-amnestic behavioral, executive, language, visuospatial, and asymmetric motor coordination phenotypes. Pathologically, AD is defined by the accumulation of β-amyloid (Aβ) plaques, hyperphosphorylated tau neurofibrillary tangles, glial changes (microglia and astrocytes), and subsequent neurodegeneration ^1,2^. Despite decades of research, its biological mechanism remains incompletely understood, making early diagnosis and effective management challenging.

The greatest known risk factor for developing AD is age followed by Apolipoprotein E (*APOE*) genetic status ^3,4^. Cardiovascular and metabolic risk factors have also been shown to affect risk in typical amnestic late-onset AD (LOAD) presentations, while novel risk factors may apply in earlier onset and non-amnestic presentations ^5^. The differential risk across AD presentations highlights our incomplete understanding of AD vulnerability. Increasingly, sex has emerged as a critical factor in understanding AD risk, progression, and clinical manifestations, as women account for approximately two-thirds of AD cases in the United States, with a higher lifetime risk of developing AD than men ^6^. Recent studies indicate that sex may modify multiple aspects of AD, including susceptibility of AD, progression, and molecular pathology including *APOE* ^7–12^. Men with AD tend to have a shorter survival time, while women experience greater memory impairment ^13–15^. Yet, here too, the mechanisms underlying these sex differences remain poorly understood, underscoring the need for more research into sex-specific risk factors associated with AD.

Recent advances in various types of big data and machine learning have enabled large-scale, data-driven investigations of AD heterogeneity. Transcriptomic approaches have identified three molecular subgroups of AD, each characterized by distinct combinations of dysregulated pathways, including susceptibility to tau-mediated neurodegeneration, amyloid-β–related neuroinflammation, synaptic signaling, immune activity, mitochondrial organization, and myelination^16^. Similarly, neuroimaging studies using MRI have revealed three neuroanatomical clusters, each defined by different initial regions of atrophy onset ^17^. Cognitive subtypes have also been identified based on performance in memory, visuospatial ability, language, and executive function. However, these studies are largely restricted to carefully selected research cohorts, and it remains unclear whether their findings generalize to larger, more diverse populations.

In contrast, electronic medical records (EMRs) capture a wide range of real-world health data, including multimorbidity patterns, medication histories, and disease trajectories, making them valuable resources for characterizing heterogeneity in AD populations ^18^. Prior EMR-based studies have identified AD subgroups associated with cardiovascular disease, psychiatric disorders, age of onset, and sensory impairments ^19^. Another study identified five distinct patient groups, characterized by mental health conditions, nontypical AD, typical AD, cardiovascular disease, and combined memory problems with cancer ^20^. From a sex-specific perspective, evidence suggests that men and women differ in comorbidity profiles ^21^ and may follow distinct AD progression trajectories ^19,22^. Similar heterogeneity in comorbidity patterns has also been reported across racial and ethnic groups ^23^. Nevertheless, subtyping methodologies are not yet standardized, with substantial variability in study populations (e.g., case definitions including early-vs. late-onset AD, and inclusion or exclusion of healthy controls) and analytic approaches. Comprehensive and unbiased analyses focusing specifically on sex-specific patterns among data-driven groupings of AD remain largely unexplored.

In this study, we apply unsupervised clustering to a large EMR dataset of AD patients, consisting largely of LOAD presentations, to investigate heterogeneity within the AD population rather than relying on traditional case-control comparisons. By leveraging real-world clinical data, we aim to characterize comorbidity patterns and examine potential sex-specific differences. We further evaluate the consistency of the identified patterns using an independent dataset. These analyses are intended to provide insights into the heterogeneity of AD and serve as a basis for hypothesis generation in future studies.

## Results

### Patient characteristics

AD cohorts were identified from the University of California (UC) San Francisco (UCSF) dataset and an independent UC-Wide dataset from which UCSF patients were excluded. This study was designed as a cross-sectional analysis at the patient level, where diagnoses across all visits were aggregated into a single binary profile for each patient to ensure independence of observations. We identified 8,804 patients who met the AD case definition in the UCSF EMRs. Of these, 441 patients (5.0%) had no recorded comorbidities beyond AD and were excluded, resulting in a final cohort of 8,363 AD patients (5,315 females (64%); median age: 90.0 [IQR: 84.0–91.0]) (Figure 1). From the UC-Wide independent dataset, we identified 25,896 AD patients (16,036 females (61.9%); median age: 89.0 [IQR: 82.0–90.0]) after applying the same inclusion and exclusion criteria. Note that age distributions among the oldest participants should be interpreted descriptively, as age was capped during de-identification through birth-date recoding in the UCSF dataset and assignment of age 91 to individuals aged 91 years or older in the UC-Wide dataset. In both datasets, AD patients were predominantly “White” (62.0% at UCSF, 65.7% in the UC-Wide dataset), followed by those classified as “Other” and “Asian”. At UCSF, most AD patients (75.0%) were recorded as “Alive,” whereas in the UC-Wide dataset, only 50.9% were alive. Additionally, the median number of comorbidities recorded at UCSF was lower compared to the UC-Wide dataset (UCSF: median = 23.0 [IQR: 9.0–63.0]; UC-Wide: median = 40.0 [IQR: 16.0–88.0]). Other demographic characteristics of AD patients are presented in Table 1 and 2.

**Figure 1.**
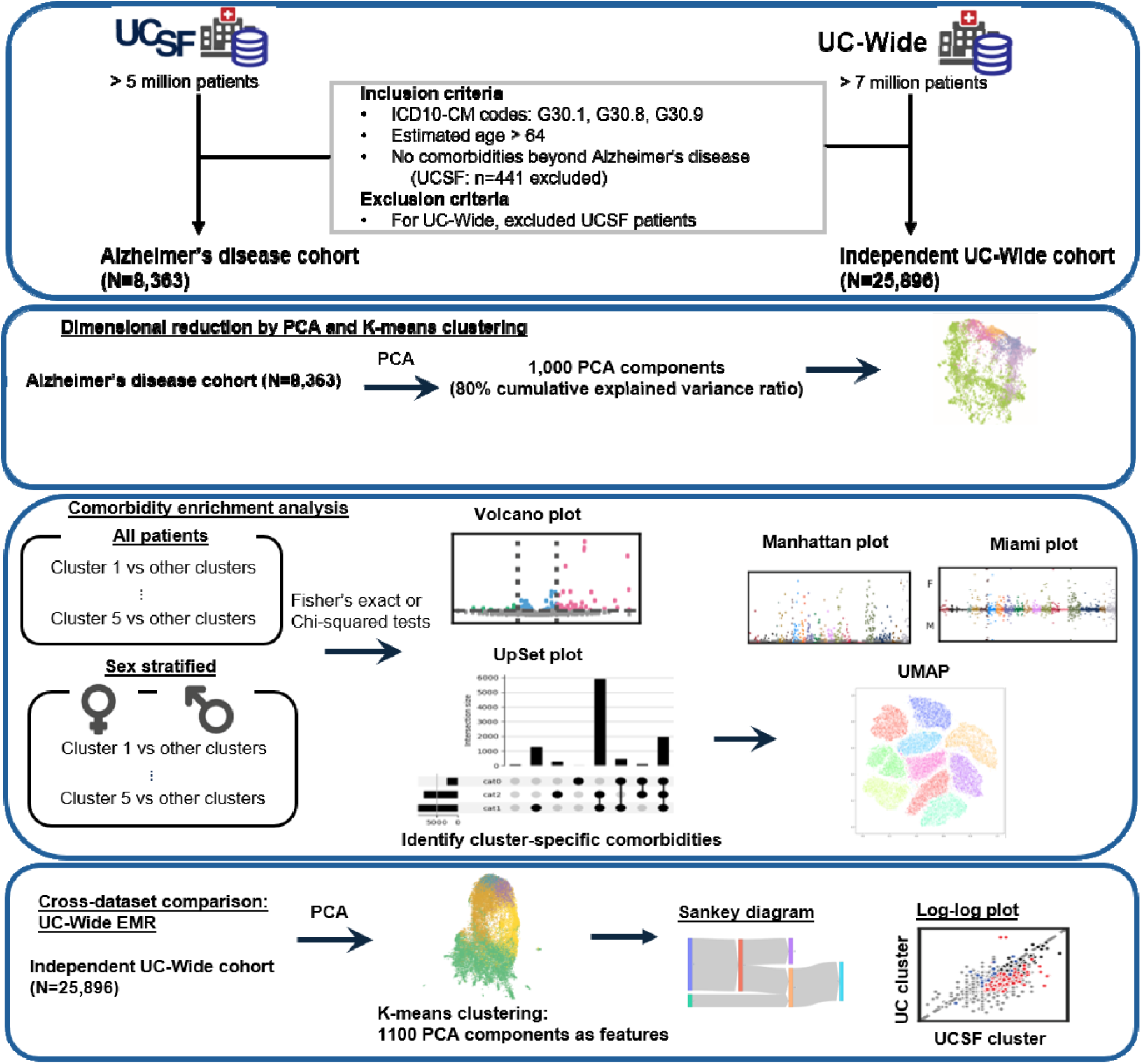
Cohort selection and overview of study design. Alzheimer’s disease patients > 64 years with at least one comorbidity were included in this study. (1) K-means clustering was performed following Principal Component Analysis (PCA). (2) Each cluster was characterized based on cluster-specific comorbidities. For external reproducibility assessment, (3) the same approach was applied to the UC-Wide cohort, excluding UCSF patients. ICD-10-CM, the International Classification of Diseases, 10th Revision, Clinical Modification; PCA, Principal Component Analysis; UC, University of California

### Low-dimensional embeddings derived from principal components of diagnostic data reveal five comorbidity patterns of AD

The analysis data consisted of patient-level diagnostic records, where each row represented an individual patient and each column corresponded to one of 33,031 unique diagnosis names across 8,363 AD patients. Diagnosis names were one-hot encoded, with a binary value indicating the presence or absence of each diagnosis for a given patient. To mitigate the impact of redundant or noisy features, Principal Component Analysis (PCA) was applied to diagnosis names (33,031 diagnosis name features) prior to K-means clustering. For the UCSF dataset, we selected 1,000 principal components, capturing approximately 80% of the cumulative explained variance, as input features for clustering.

Cluster selection was guided by a combination of quantitative metrics and interpretability, including within-cluster sum of squares (WSS), silhouette scores, Calinski–Harabasz (CH) index, and Davies– Bouldin (DB) index. Across these metrics, solutions with two or three clusters yielded more compact and well-separated partitions. However, these solutions resulted in overly coarse groupings that failed to capture clinically meaningful heterogeneity. Increasing the number of clusters from 4 to 5 resulted in only a modest improvement in within-cluster homogeneity (ΔWSS = 0.0077), while maintaining a reasonable cluster size distribution (smallest cluster = 3.9%). In contrast, increasing the number of clusters to 6 resulted in minimal improvement in WSS (ΔWSS = 0.004), a decrease in silhouette score (0.019), and an increase in the DB index, suggesting over-segmentation and reduced clustering quality (Table S1).

Although the improvement in quantitative metrics from k = 4 to k = 5 was limited, examination of co-occurring diagnosis pairs based on their prevalence suggested that the additional cluster allowed further separation of previously heterogeneous patterns. Specifically, most patients assigned to Cluster 2 in the four-cluster solution were redistributed into Clusters 2 and 4 in the five-cluster solution. In the four-cluster solution, Cluster 2 represented a heterogeneous population characterized by hypertension and a mixture of symptoms and comorbid conditions. In contrast, the five-cluster solution separated this group into two clusters: Cluster 2, reflecting more general or milder conditions and healthcare utilization, and Cluster 4, enriched for more clinically relevant comorbidities, including infections and hematologic conditions. Taken together, the five-cluster solution was selected as an interpretability-informed representation of the dataset that provided additional granularity compared with smaller cluster solutions. However, alternative solutions with fewer clusters were also supported by all clustering indices (Table S2, Data S1).

We characterized each cluster based on comorbidity profiles. In this step, comorbidities were defined using International Classification of Diseases, 10th Revision, Clinical Modification (ICD-10-CM) codes to facilitate clinical interpretation and enhance the interpretability of cluster characteristics. Within the UCSF clusters, Cluster 5 had significantly fewer comorbidities (median = 12.0 [IQR: 5.0–21.0], Bonferroni-corrected p-values < 0.05) compared to the other clusters, whereas Cluster 1 had the highest number of comorbidities (median = 313.5 [IQR: 258.0–379.0], Bonferroni-corrected p-values < 0.05). In terms of age distribution, Cluster 4 (median = 91.0 [IQR: 90.0–91.0], Bonferroni-corrected p-values < 0.05) was significantly older than Cluster 1, 3, and 5, followed by Cluster 2. Conversely, Cluster 3 (median = 88.0 [IQR: 82.0–90.0], Bonferroni-corrected p-values < 0.05) was significantly younger than all other clusters (Table S3). Black or African American individuals comprised 13% of Cluster 2 and 12% of Cluster 4, representing a higher proportion compared to other clusters (Bonferroni-corrected p-values < 0.05, Table S3). Regarding sex, Cluster 5 included a significantly higher proportion of males than Cluster 4, with these males displaying a younger age distribution. Other demographic characteristics and sex-stratified distributions for each cluster are presented in Table 1 and Table S3.

For the UC-Wide dataset, 19,835 unique diagnosis names from 25,896 AD patients were used as features for PCA. 1,100 principal components covering approximately 80% of the cumulative explained variance were selected as input features for K-means clustering. Consistent with the UCSF dataset, clustering solutions with smaller numbers of clusters (k=2–3) yielded higher silhouette and CH scores, indicating more compact and well-separated partitions. However, these solutions resulted in overly coarse groupings that failed to capture clinically meaningful heterogeneity in comorbidity patterns. As the number of clusters increased, improvements in within-cluster homogeneity diminished progressively. Increasing the number of clusters from 5 to 6 resulted in minimal reduction in WSS (ΔWSS = 0.6%), accompanied by a further decline in silhouette score and the emergence of a smaller cluster (3.7% of the population), indicating the onset of over-segmentation. Taken together, k=5 was selected as the highest-resolution partition that balances cluster interpretability, granularity, and parsimony, while avoiding over-fragmentation (Table S1).

Significant differences in the number of comorbidities were observed between Cluster A (median = 201.0 [IQR: 166.0–253.0], Bonferroni-corrected p-values < 0.05) and Cluster B (median = 15.0 [IQR: 7.0–25.0], Bonferroni-corrected p-values < 0.05) (Table S3). Other demographic characteristics of each cluster are presented in Table 2. A low-dimensional Uniform Manifold Approximation and Projection (UMAP) visualization of the PCA-transformed components illustrates the distribution of AD patient clusters, sex differences, and the number of comorbidities (Figure 2).

**Figure 2.**
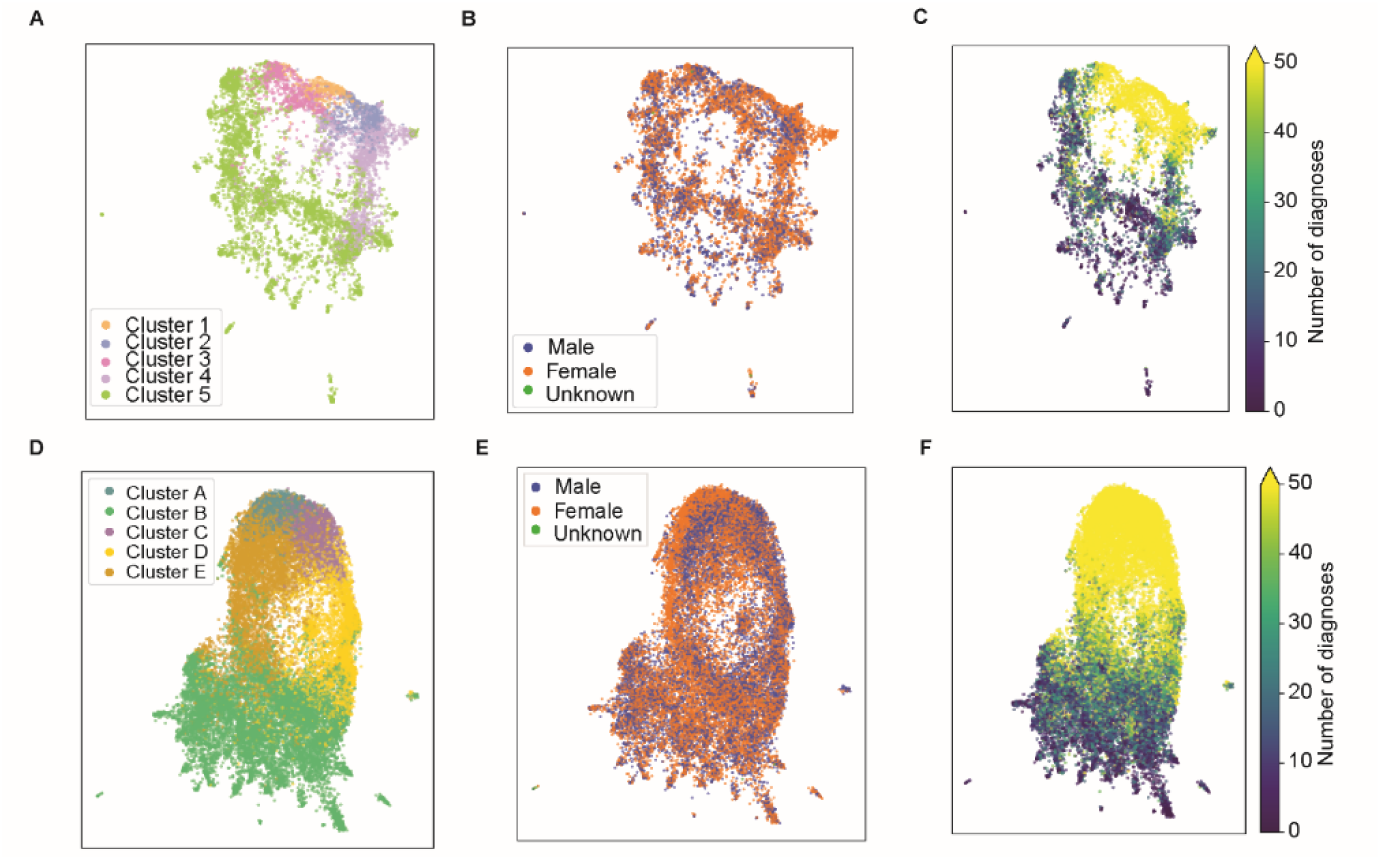
UMAP visualizations using PCA components as features. UMAP plots are colored by cluster (**A** and **D**), sex (**B** and **E**), and number of comorbidities (**C** and **F**). Each dot represents a patient. **A–C** correspond to the UCSF cohort, while **D–F** represent the independent cohort in the UC-Wide dataset. UC, University of California

### Comorbidity enrichment analysis shows significant cluster-related comorbidities at UCSF

Our analysis identified significant diagnoses in each cluster within the UCSF dataset (two-sided Fisher’s exact or Chi-squared test, Bonferroni-corrected p-value < 0.05) (Figure 3). Cluster 1 had 1,911 significant comorbidities, most of which were positively associated. In contrast, 1,650 significant comorbidities were identified in Cluster 5, predominantly showing negative associations. However, a few conditions exhibited positive associations, including Pick’s disease. Cluster 2 – 4 displayed unique comorbidity enrichment patterns. For example, in Cluster 2 (985 significant comorbidities), the top positively associated diagnoses included laceration, phlebitis and thrombophlebitis of femoral vein, and benign neoplasm of the conjunctiva. Interestingly, Cluster 3 (682 significant comorbidities) showed relatively lower prevalence of mental disorders and personality changes. Finally, Cluster 4 (245 significant comorbidities) exhibited positive associations with pneumonia and disseminated intravascular coagulation (Data S1). Manhattan plots illustrated the distribution of statistical significance (Figure S1). However, although many comorbidities showed statistically significant associations, their relatively low prevalence may limit their clinical relevance, and these findings should therefore be interpreted with caution.

**Figure 3.**
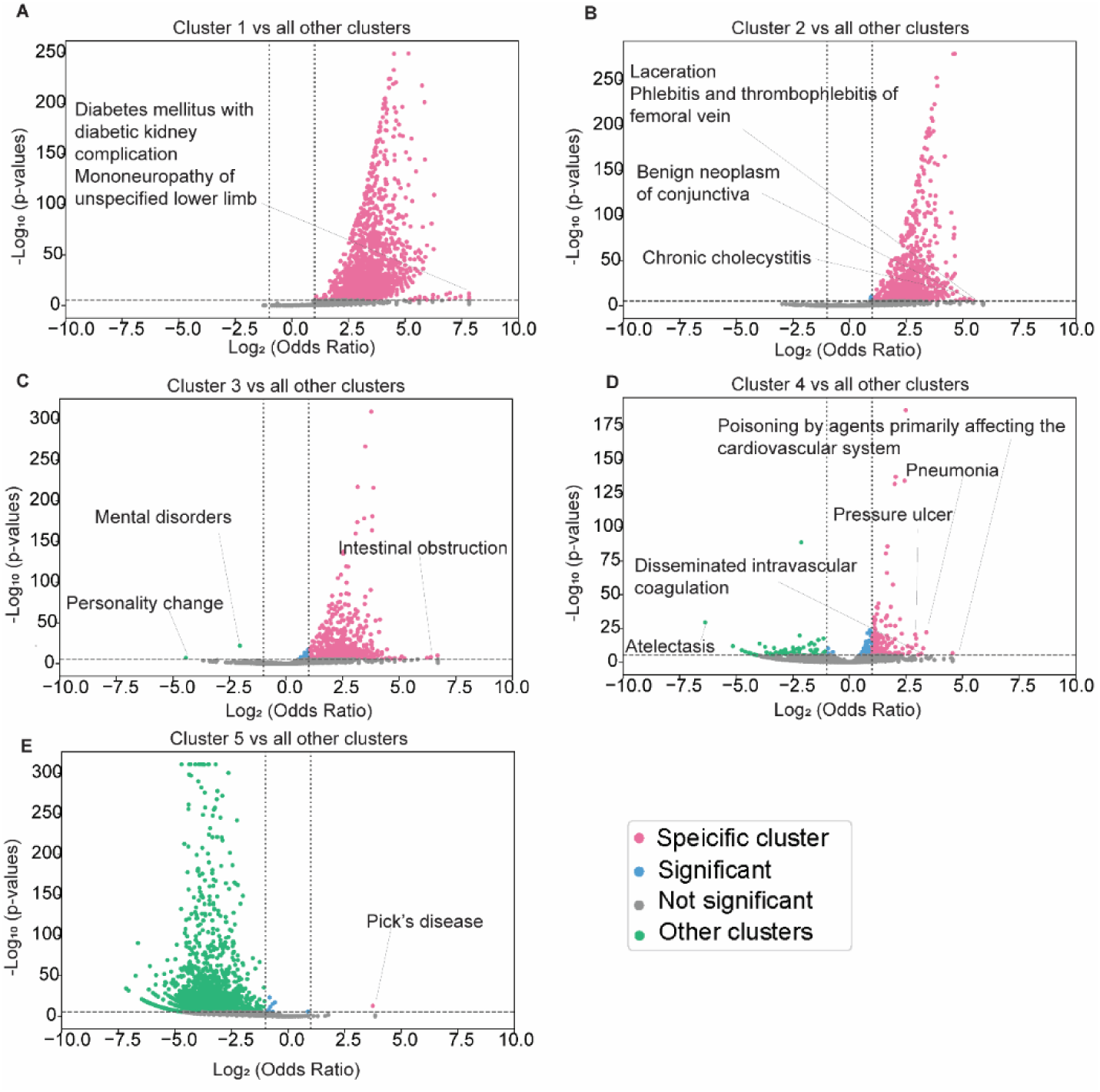
Volcano plots of cluster-specific comorbidity enrichment patterns. Volcano plots display enriched ICD-10-CM codes identified using the two-sided Fisher’s exact test (for cases < 5) or the Chi-squared test (for cases ≥ 5). Enrichment was determined based on a Bonferroni-corrected p-value < 0.05. Odds ratios were calculated by comparing each specific cluster to all other clusters. **A:** Cluster 1, **B:** Cluster 2, **C:** Cluster 3, **D:** Cluster 4, **E:** Cluster 5. ICD-10-CM, the International Classification of Diseases, 10th Revision, Clinical Modification.

### Significant cluster-specific comorbidities at UCSF showed partial consistency in the UC-Wide independent cohort

We identified cluster-specific comorbidities using UpSet plots (Figure 4). Cluster 1 exhibited the highest number of positively associated comorbidities (812 comorbidities), followed by Cluster 2 (225 comorbidities). Cluster 3 had 96 positively associated comorbidities and one negatively associated comorbidity (personality change), while Cluster 4 had 13 positively and 6 negatively associated comorbidities. In contrast, Cluster 5 had only two positively associated comorbidities, whereas 1,581 comorbidities were negatively associated. Examining the most significantly associated comorbidities specific to each cluster, we found that Cluster 3 was characterized by acute systolic heart failure and ileus, whereas pneumonia and brief psychotic disorder were observed in Cluster 4. In Cluster 5, we observed essential hypertension, anemia, and urinary tract infection as the most significantly negatively associated comorbidities (Figure 5).

**Figure 4.**
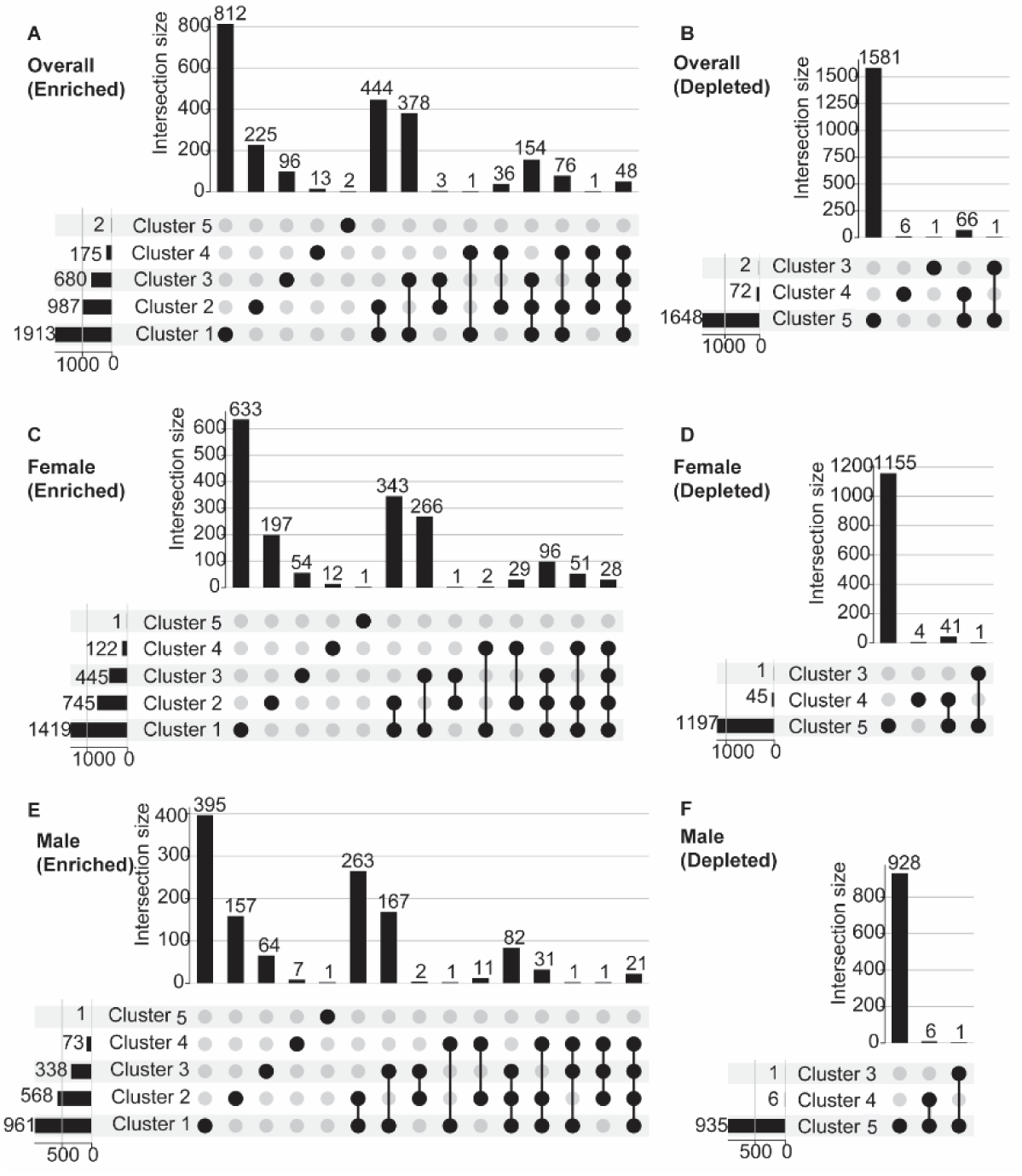
UpSet plots of cluster-specific comorbidity enrichment patterns. UpSet plots illustrating significant cluster-specific comorbidities. Panels represent the overall population (**A:** Positive associations, **B:** Negative associations), females (**C:** Positive associations, **D:** Negative associations), and males (**E:** Positive associations, **F:** Negative associations).

**Figure 5.**
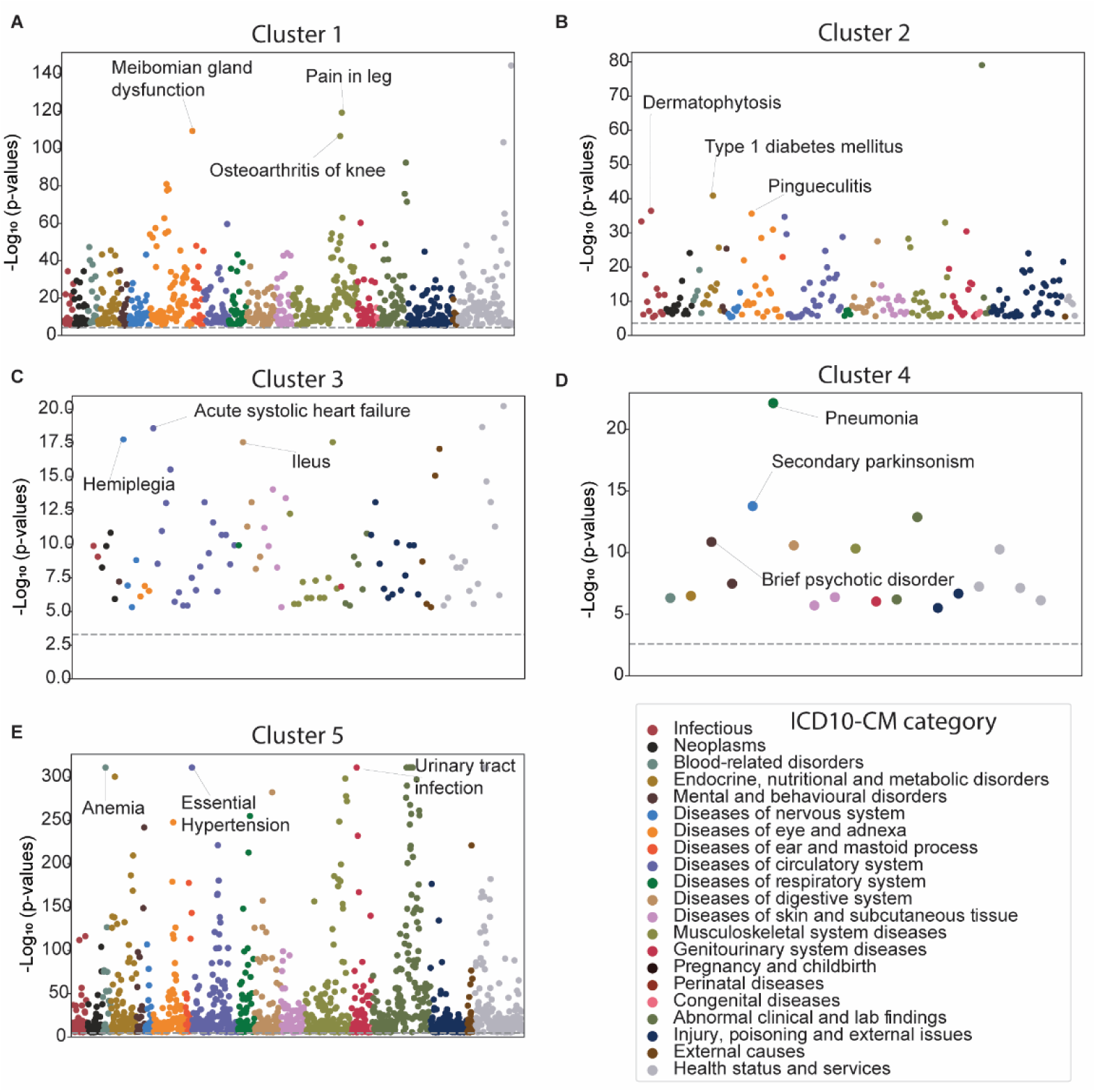
Manhattan plots of cluster-specific comorbidity enrichment patterns. Manhattan plots displaying enriched ICD-10-CM codes identified using the two-sided Fisher’s exact test (for cases < 5) or the Chi-squared test (for cases ≥ 5). Enrichment was determined based on a Bonferroni-corrected p-value < 0.05. **A:** Cluster 1, **B:** Cluster 2, **C:** Cluster 3, **D:** Cluster 4, **E:** Cluster 5. ICD-10-CM, the International Classification of Diseases, 10th Revision, Clinical Modification.

Additionally, cluster-specific comorbidities with high odds ratios were identified (Figure 6A). Because many of these diagnoses were uncommon, they should be interpreted as low-frequency signals rather than defining clinical features of a cluster.

**Figure 6.**
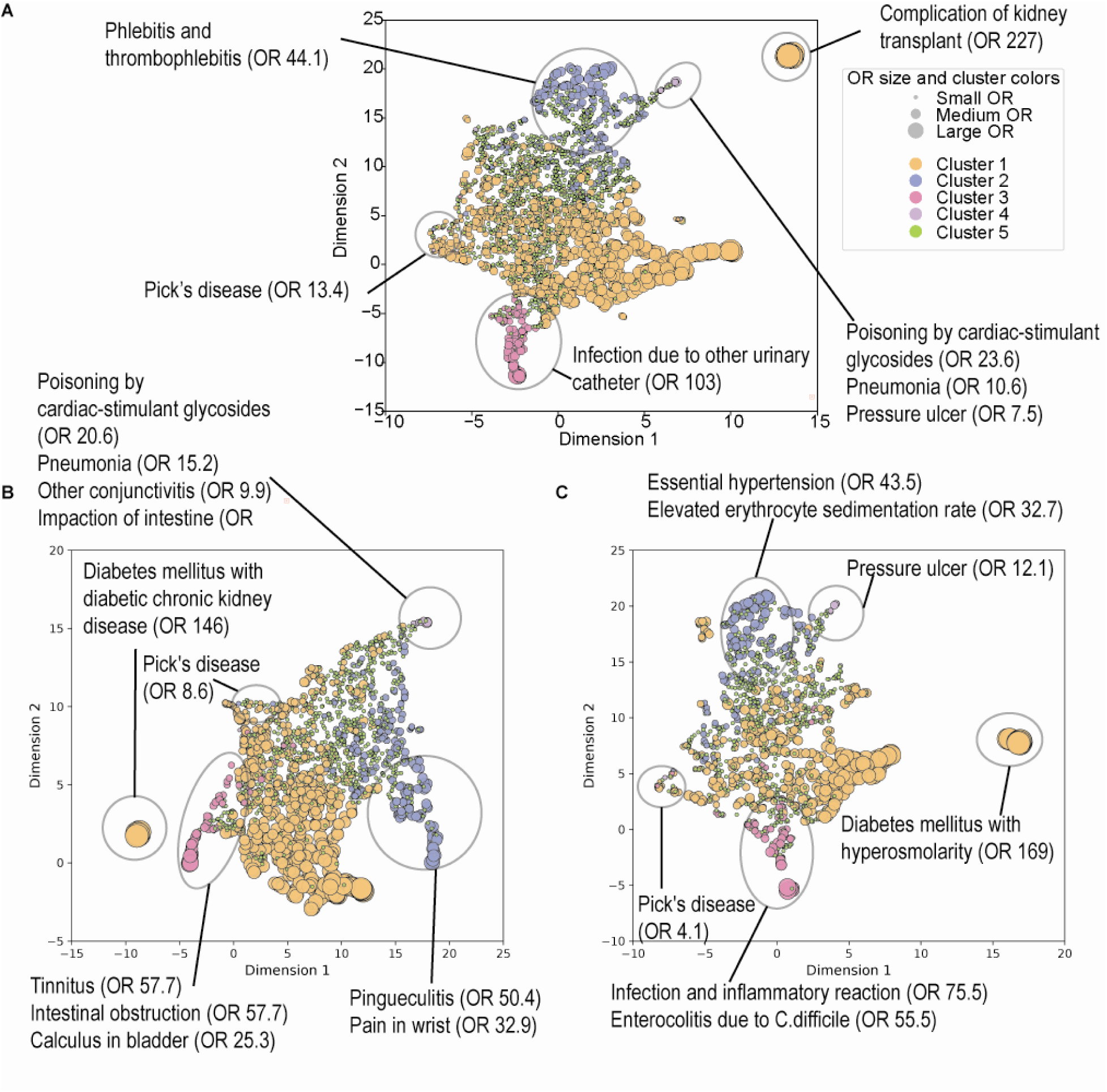
UMAP visualizations of cluster-specific comorbidities using odds ratios as features. This figure presents a UMAP visualization of ICD-10-CM codes based on their standardized ORs across different clusters. Each dot represents a specific ICD-10-CM code, with the dot size corresponding to the OR of the significantly associated cluster and the color indicating the corresponding cluster. The input features for the UMAP projection were derived from a matrix of standardized ORs of cluster-specific comorbidities. These standardized ORs were calculated by comparing the prevalence of each diagnosis within a specific cluster against all other clusters using Fisher’s exact test (for cases < 5) or the Chi-squared test (for cases ≥ 5). A Bonferroni-corrected p-value threshold of 0.05 was applied to identify significantly enriched diagnoses. **A:** Overall population, **B:** Female, **C:** Male. ICD-10-CM, International Classification of Diseases, 10th Revision, Clinical Modification; UMAP, Uniform Manifold Approximation and Projection.

Cluster 1 showed numerous significant associations with a variety of conditions, including diabetes mellitus with other diabetic kidney complication, contusion of the right upper arm, displaced fracture of shaft of right clavicle, complication of kidney transplant, encounter for aftercare following kidney transplant, and mononeuropathy of right lower limb (all ORs = 227, Bonferroni-corrected p-values < 0.05). In Cluster 2, phlebitis and thrombophlebitis of femoral vein had the highest odds ratio (OR = 44.1, Bonferroni-corrected p-value < 0.05). Cluster 3 was characterized by infection and inflammatory reaction due to a urinary catheter (OR = 103, Bonferroni-corrected p-value < 0.05), followed by intestinal obstruction (OR = 84.3, Bonferroni-corrected p-value < 0.05). Additionally, dysphagia was significantly associated with Cluster 3 (OR = 5.3, Bonferroni-corrected p-value < 0.05, Data S1). Cluster 4 was significantly associated with poisoning by cardiac-stimulant glycosides (OR = 23.6, Bonferroni-corrected p-value < 0.05), pneumonia (OR = 10.6, Bonferroni-corrected p-value < 0.05), and pressure ulcer (OR = 7.5, Bonferroni-corrected p-value < 0.05). Lastly, in Cluster 5, Pick’s disease was significantly enriched (OR = 13.4, Bonferroni-corrected p-value < 0.05). However, odds ratios in our study should be interpreted with caution, as the frequency of certain diagnoses in each cluster was relatively small, which may have led to inflated odds ratios (Data S1). Indeed, a substantial proportion of cluster-specific comorbidities had a prevalence below 5%, suggesting that these findings are better interpreted as statistically enriched but low-frequency signals.

To improve clinical interpretability and reduce the influence of sparse diagnoses with inflated odds ratios, we also examined more common comorbidities (prevalence ≥5%). Cluster 1 showed patterns consistent with age-related multimorbidity, including ophthalmologic, musculoskeletal, and vascular conditions. Cluster 2 showed enrichment for cardiometabolic and cerebrovascular conditions, including diabetes, cerebral infarction, and heart failure. In contrast, Cluster 5 showed negative associations with common chronic conditions (e.g., hypertension, hyperlipidemia). Clusters 3 and 4 were not well characterized by common conditions, as the majority of associated comorbidities were of low prevalence (Data S1).

In the UC-Wide independent dataset, we identified corresponding UC-Wide clusters for each UCSF cluster based on the overlap in significant comorbidities, as visualized by Sankey plots (Figure 7A). The proportion of significant comorbidities in each UCSF cluster that were also captured in the corresponding UC-Wide clusters ranged from 20% to 55%, indicating partial overlap across datasets. For instance, 239 of the 812 (29.4%) significant comorbidities identified in UCSF Cluster 1 were also observed in the matched UC-Wide Cluster A, which itself contained 1,976 cluster-specific significant comorbidities.

**Figure 7.**
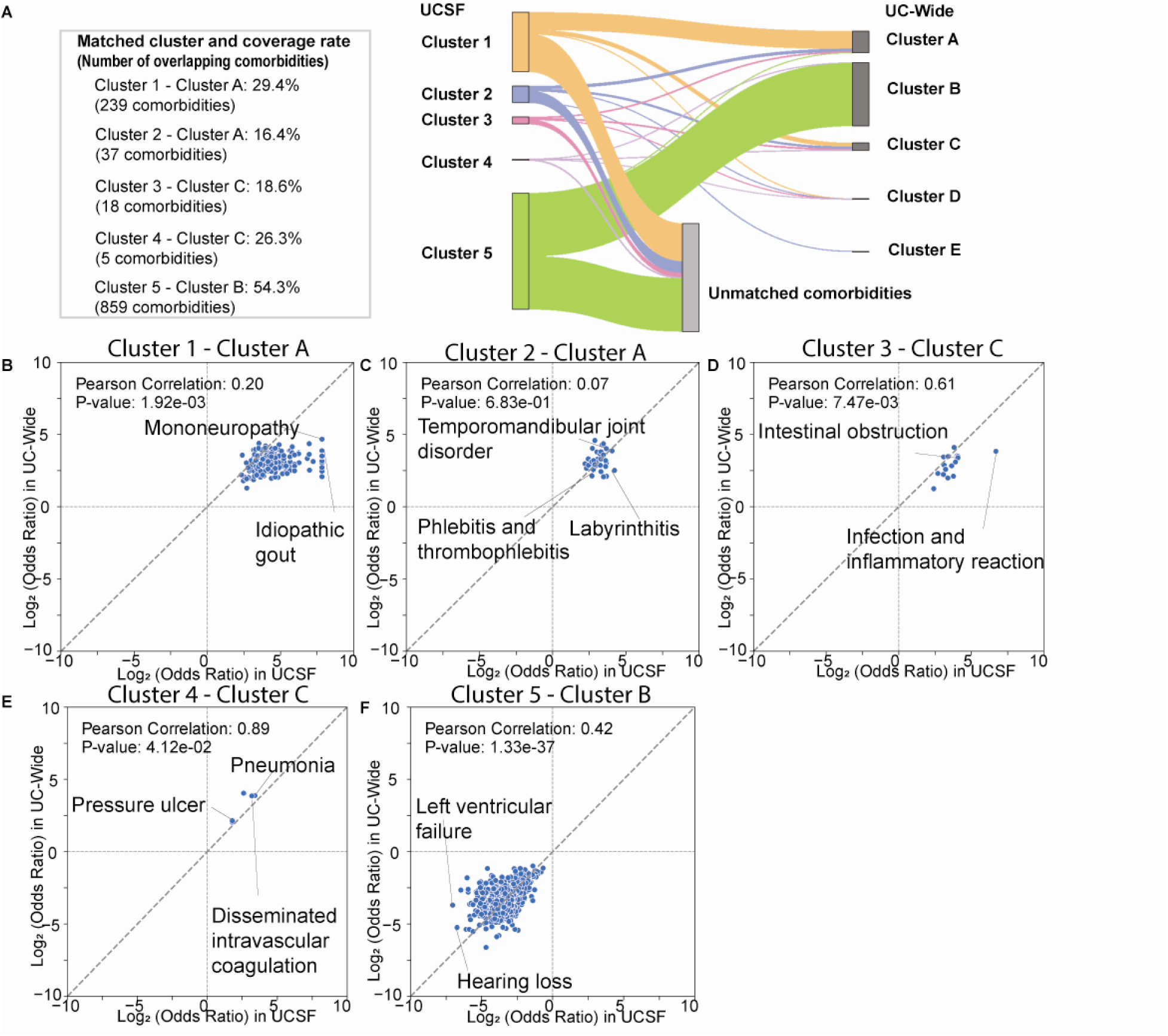
Cross-dataset comparison of cluster-specific comorbidities. **A** Sankey plot illustrating the overlap of significant comorbidities across UCSF and UC-Wide populations within each cluster. The coverage rate represents the proportion of UCSF cluster-specific comorbidities that remained significant in the matched UC-Wide cluster. Number of cluster-specific comorbidities: Cluster 1 (812), Cluster 2 (225), Cluster 3 (97), Cluster 4 (19), Cluster 5 (1,583), Cluster A (1,976), Cluster B (2,731), Cluster C (819), Cluster D (40), Cluster E (19) **B–F** Log-log plots showing partial reproducibility of comorbidity enrichment patterns across datasets. **B:** Cluster 1 vs. Matched UC-Wide Cluster A, **C:** Cluster 2 vs. Matched UC-Wide Cluster A, **D:** Cluster 3 vs. Matched UC-Wide Cluster C, **E:** Cluster 4 vs. Matched UC-Wide Cluster C, **F:** Cluster 5 vs. Matched UC-Wide Cluster B P-values were calculated from Pearson correlation analysis. UC, University of California.

To further evaluate the consistency of comorbidity enrichment across datasets, we examined the correlation of shared comorbidities using Log-Log plots (Figure 7B). Partial consistency was observed for several UCSF cluster-specific comorbidities with high odds ratios, including phlebitis and thrombophlebitis in Cluster 2, intestinal obstruction, and infection and inflammatory reaction due to a urinary catheter in Cluster 3, and pneumonia and pressure ulcers in Cluster 4. However, the overall consistency across clusters was limited. Notably, Cluster 2 did not show a significant correlation with its matched UC-Wide Cluster A (Pearson r = 0.07, p = 0.68). In contrast, clusters representing broader comorbidity patterns, particularly Cluster 1 and Cluster 5, showed relatively more consistent patterns across datasets. Consistency in comorbidity prevalence across datasets was limited, with no comorbidities (prevalence ≥5%) consistently observed in Clusters 1–4, whereas broader patterns in Cluster 5 showed partial overlap (Figure S2).

### Sex-stratified analysis identifies significantly associated cluster-sex-specific comorbidities at UCSF, some of which show partial consistency in the UC-Wide independent cohort

After illustrating the distribution of statistical significance in Miami plots, we identified cluster-sex-specific comorbidities using UpSet plots (Figure 4, Figure S3). When we focus on the significant comorbidities exclusive to each cluster, in Cluster 1, females showed a strong association with pain-related conditions, whereas males were primarily associated with symptoms of the genitourinary system. In Cluster 2, females exhibit a strong association with tension-type headaches, whereas males are more frequently associated with circulatory diseases, including unstable angina and hypertensive heart disease. In Cluster 3, females show significant associations with pressure ulcers and osteoarthritis, while males are predominantly linked to respiratory diseases, such as acute pulmonary edema, and circulatory conditions, including acute systolic heart failure. In Cluster 4, females are significantly associated with pneumonia. In Cluster 5, we observed essential hypertension, anemia, and urinary tract infection across both sexes (Figure 8).

**Figure 8.**
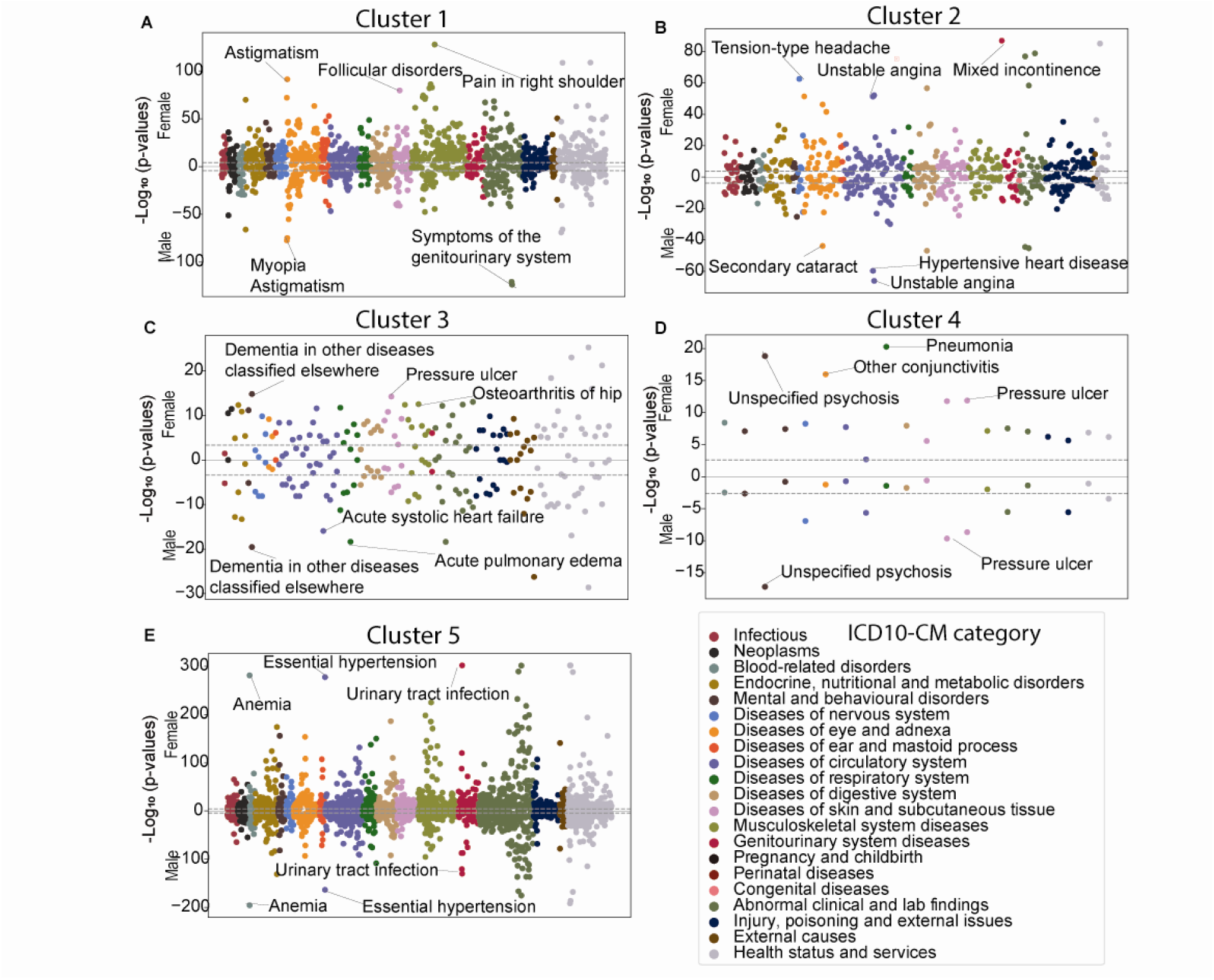
Miami plots of cluster-specific comorbidity enrichment patterns. Miami plots displaying enriched ICD-10-CM codes identified using the two-sided Fisher’s exact test (for cases < 5) or the Chi-squared test (for cases ≥ 5). Enrichment was determined based on a Bonferroni-corrected p-value < 0.05. ICD-10-CM, the International Classification of Diseases, 10th Revision, Clinical Modification. **A:** Cluster 1, **B:** Cluster 2, **C:** Cluster 3, **D:** Cluster 4, **E:** Cluster 5.

Considering odds ratios, diabetes-related conditions were significantly associated with Cluster 1 in both sexes, with diabetes mellitus due to an underlying condition with diabetic chronic kidney disease being predominant in females (OR = 146) and diabetes mellitus with hyperosmolarity in males (OR = 169). Similarly, Pick’s disease remained highly associated with Cluster 5 across both sexes (Female OR = 8.6, Male OR = 4.1). In Cluster 2, females exhibit stronger associations with eye diseases, such as pingueculitis (OR=50.4, Bonferroni-corrected p-value < 0.05), whereas males are more associated with circulatory diseases, including hypertension (OR=43.5, Bonferroni-corrected p-value < 0.05) and elevated erythrocyte sedimentation rate (ESR) (OR=32.7, Bonferroni-corrected p-value < 0.05). In Cluster 3, females demonstrate high odds ratios for tinnitus (OR=57.7, Bonferroni-corrected p-value < 0.05), intestinal obstruction (OR=57.7, Bonferroni-corrected p-value < 0.05), and bladder calculus (OR=25.3, Bonferroni-corrected p-value < 0.05), while males are significantly associated with infection and inflammatory reaction (OR=75.5, Bonferroni-corrected p-value < 0.05) and enterocolitis (OR=55.5, Bonferroni-corrected p-value < 0.05). Lastly, in Cluster 4, females have strong associations with poisoning by cardiac-stimulant glycosides (OR=20.6, Bonferroni-corrected p-value < 0.05) and pneumonia (OR=15.2, Bonferroni-corrected p-value < 0.05), whereas males are associated with pressure ulcers (OR=12.1, Bonferroni-corrected p-value < 0.05) (Figure 6B and 6C, Data S1).

To aid clinical interpretation, common comorbidities (prevalence ≥5%) were also considered. Under this perspective, no substantial sex differences were observed in Clusters 1, 3, and 5. Cluster 2 showed a more pronounced pattern of cardiovascular-related conditions among males, whereas in Cluster 4, the male subgroup was enriched for conditions such as secondary Parkinsonism and neuropsychiatric disorders (Data S1). However, these findings should be interpreted with caution due to the limited number of cases for certain diagnoses, which may lead to variability in estimates.

In the UC-Wide dataset, we identified corresponding UC-Wide clusters based on the coverage rate of UCSF cluster-specific comorbidities in each sex (Figure S4). The matched UC-Wide clusters remained consistent with those identified in the overall population, except for UCSF Cluster 3 among females. For UCSF Cluster 4 among males, two UC-Wide clusters had the same coverage rate. In this case, the cluster that was consistently matched in the overall analysis and had one of the highest coverage rates was selected as the matched cluster. Cross-dataset consistency was observed for some UCSF-derived comorbidity patterns, although reproducibility varied across clusters. Overall, partial cross-dataset consistency was observed for several UCSF cluster-specific comorbidities with high odds ratios, including elevated ESR in Cluster 2 among males, intestinal obstruction in Cluster 3 among females, and infection and inflammatory reaction due to a urinary catheter and enterocolitis in Cluster 3 among males. However, the overall consistency across clusters was limited, and some Pearson correlation coefficients were neither strong nor statistically significant (Figure 9). Cross-dataset consistency in comorbidity prevalence was also limited. Comorbidities with prevalence ≥5% in Clusters 1 and 5 partially overlapped across datasets, whereas more cluster-specific or lower-prevalence signals showed limited reproducibility in the independent UC-Wide dataset (Figure S5).

**Figure 9.**
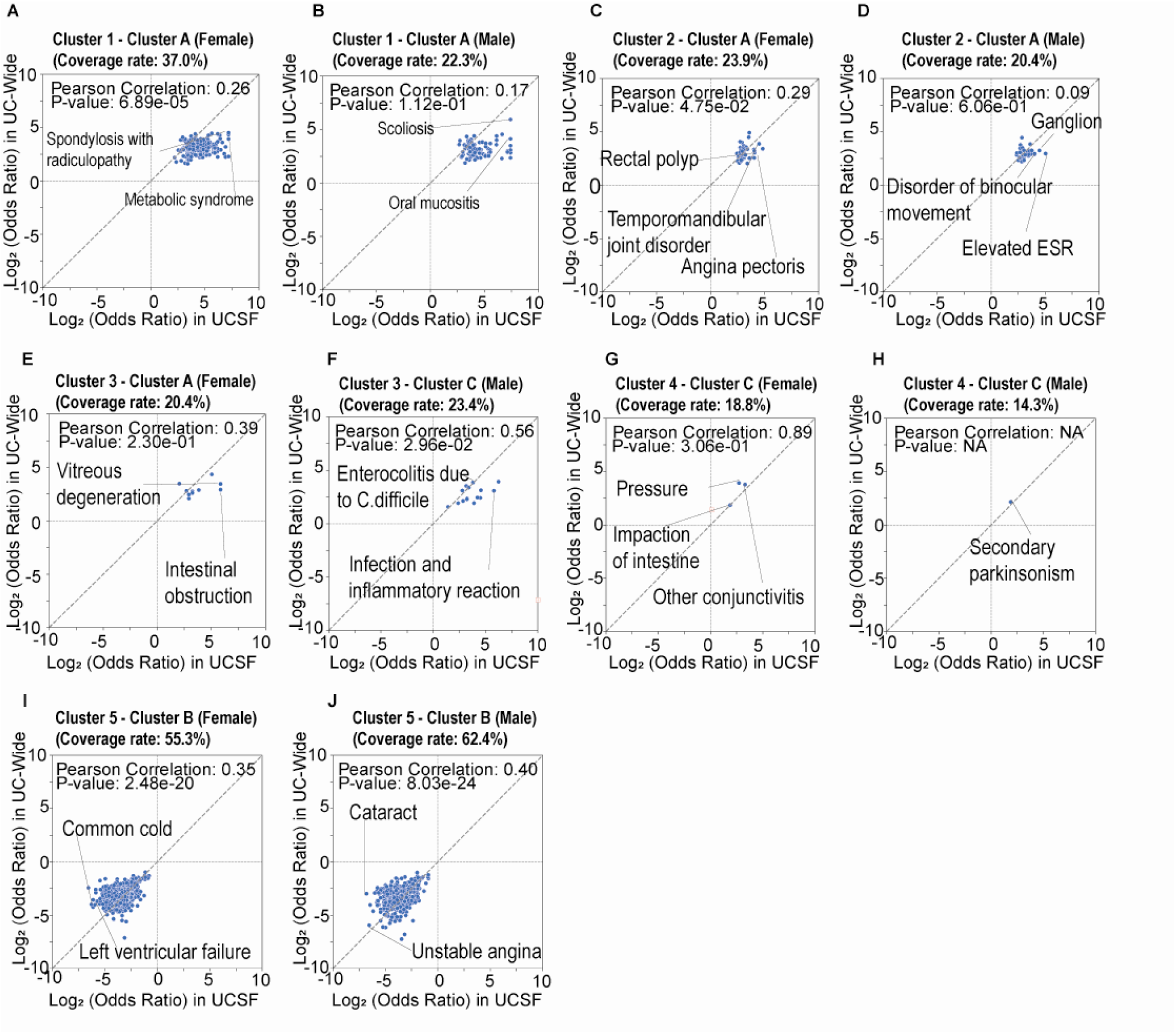
Cross-dataset comparison of cluster-specific comorbidities across sex. Log-log plots illustrating partial reproducibility of comorbidity enrichment patterns across datasets in sex-stratified populations. The coverage rate represents the proportion of UCSF cluster-sex-specific comorbidities that remained significant in the matched UC-Wide cluster. **A, C, E, G, I**: Cluster 1–Cluster 5 (Female) vs. Matched UC-Wide Cluster (Female), **B, D, F, H, J:** Cluster 1–Cluster 5 (Male) vs. Matched UC-Wide Cluster (Male). P-values were calculated from Pearson correlation analysis. UC, University of California.

## Discussion

In this study of 8,363 AD patients from UCSF and 25,896 from an independent UC-Wide cohort, we analyzed multi-site EMR by applying dimensionality reduction, followed by clustering analysis, and in the process, we identified five dataset-derived comorbidity clusters among AD patients.

A key strength of our approach is its focus on patients with LOAD, enabling the characterization of heterogeneity within the most common clinical presentations and minimizing potential noise from other dementia types. Importantly, whereas most prior studies have compared AD patients with cognitively normal controls^19,21,23^, our study advances the field by performing comparisons within the AD population, thereby providing a structured characterization of heterogeneity among patients. We further characterized comorbidity profiles by integrating statistical association (odds ratios) with clinical relevance (prevalence), using odds ratios to identify potentially informative signals and prevalence to contextualize their clinical importance. Finally, this study provides a systematic, data-driven characterization of comorbidity patterns within AD and evaluates their reproducibility across an independent dataset.

Several of the identified clusters corresponded to recognizable clinical syndromes, including patterns characterized by multimorbidity, cardiovascular disease burden, frailty-related conditions, gastrointestinal disorders, and urinary complications. Therefore, these findings should not be interpreted as the discovery of previously unrecognized disease entities. Rather, the contribution of this study lies in providing a structured, data-driven characterization of how these comorbidity patterns are distributed within an AD population, integrating statistical enrichment with prevalence-based interpretation, and evaluating their consistency across independent real-world datasets.

Enrichment analysis revealed clinical heterogeneity, and sex-stratified analysis highlighted potential sex-specific differences. While clusters characterized by either high overall comorbidity burden or relatively low burden showed some consistency across datasets, more cluster-specific or lower-prevalence signals showed limited reproducibility, suggesting limited generalizability of finer-grained patterns.

These findings suggest that the identified clusters are best interpreted as data-driven groupings that capture dominant patterns in the dataset. All clustering indices supported solutions with fewer clusters. Therefore, the five-cluster solution should not be interpreted as a uniquely optimal or biologically meaningful classification of AD. Rather, the five clusters represent an interpretability-informed partition of comorbidity patterns within this dataset that provides additional granularity for exploring heterogeneity. In the following sections, we present our findings for each cluster, followed by the identification of potential sex-specific differences, and discuss their implications.

### Cluster 1: high comorbidity burden with complex clinical profiles

Cluster 1 was characterized by a high comorbidity burden, which may reflect the presence of patients with complex clinical profiles. In particular, diseases with the highest odds ratios included several kidney-related conditions—diabetes mellitus with other diabetic kidney complications, complications of kidney transplant, and encounter for aftercare following kidney transplant. These findings may be associated with a complex clinical profile in this cluster. However, additional enriched conditions were also observed, reflecting a broader range of systemic complications beyond kidney disease alone. Kidney transplantation is a recognized risk factor for AD and is often associated with multiple comorbidities due to long-term immunosuppressive therapy and underlying chronic conditions^24,25^. In this context, kidney-related conditions may represent one component of a broader pattern of multimorbidity within this cluster, rather than a defining feature. Further stratification of this cluster may help identify more specific comorbidity patterns and support hypothesis-generating insights for future studies.

### Cluster 2: African American individuals, phlebitis and cardiovascular conditions

Cluster 2 presented with a relatively higher proportion of Black or African American individuals. Patients exhibited enriched comorbidities such as phlebitis and thrombophlebitis of femoral vein. While a relatively higher prevalence of cardiovascular conditions was observed in this cluster, these patterns showed limited reproducibility in the independent dataset. Therefore, the observed associations between AD and vascular abnormalities should be interpreted as exploratory.

### Cluster 3: lower rates of mental illness and increased infections and gastrointestinal dysfunction

This group exhibited significant negative associations with mental disorders. Given that mental disorders are commonly reported comorbidities in AD patients ^26^, this finding is of interest. One possible explanation for the lower prevalence of mental disorders is the significantly younger age of patients in this cluster compared to others. These findings may reflect a relatively younger subgroup within the late-onset AD (LOAD).

Additionally, we observed infection and inflammatory reactions due to a urinary catheter, as well as intestinal obstruction, as positively associated comorbidities in Cluster 3. Previous studies have reported associations of AD with gastrointestinal dysfunction^27–29^ and infection-related diagnoses^30–32^. However, the mechanisms underlying these observations could not be evaluated in the present study. Given the relatively low prevalence of some identified comorbidities, the observational nature of this study, and the lack of consistent reproducibility in the independent dataset, these findings should be interpreted cautiously as hypothesis-generating rather than indicative of specific biological mechanisms.

### Cluster 4: oldest age and enrichment of pneumonia and pressure ulcers

Patients in Cluster 4 constituted the oldest subgroup and exhibited higher odds ratios for pneumonia and pressure ulcers. These conditions are commonly observed among older individuals and may reflect increased medical complexity and functional impairment. Although both pneumonia and pressure ulcers were also observed in the corresponding cluster of the independent dataset, their prevalence was low in both datasets. Therefore, these findings should be interpreted cautiously as statistically enriched but low-frequency comorbidities rather than dominant clinical features of the cluster.

### Cluster 5: male sex, low comorbidity burden, potential overlap with frontotemporal dementia

Cluster 5 included a significantly higher proportion of males with a relatively younger age distribution. Given that AD possesses a female predominance, the enrichment of younger males is particularly intriguing. This cluster exhibited a relatively low burden of common chronic comorbidities, such as hypertension and hyperlipidemia, compared to other clusters.

While Cluster 5 showed the fewest positively associated comorbidities, there was one notable exception, Pick’s disease. Pick’s disease, also referred to as behavioral variant frontotemporal dementia (bvFTD), is second to AD in being the most common neurodegenerative disorder in individuals with first symptoms younger than 65 ^33^ and is known to have a higher prevalence of male sex ^34,35^. Considering the demographics of Cluster 5, this pattern may suggest some overlap with conditions that can share clinical features with AD.

However, the Pick’s disease signal did not show consistent reproducibility in the independent dataset, and given the observational nature of this study and the reliance on EHR-based diagnostic coding, this finding should be interpreted as hypothesis-generating. Future studies integrating imaging or biomarker data would be valuable to determine whether this cluster captures patients with bvFTD or other diagnostic entities.

### Sex specific analysis

While sex related results only came out in our first pass analysis in cluster 5, given the significant female bias to AD, we performed follow up sex specific analyses. These analyses may provide preliminary insights into sex-associated comorbidity patterns observed in diagnostic coding data, particularly in Clusters 2 and 3.

In Cluster 2, hypertension and elevated ESR were more frequently observed in males. Our findings may reflect a vascular and metabolic comorbidity pattern in this subgroup. Differences in demographic composition between male and female subgroups may partially contribute to this pattern. In particular, the male subgroup had a higher proportion of White individuals and a lower proportion of Black individuals compared to the female subgroup.

In Cluster 3, our study observed that calculus in the bladder was associated with females. Although this observation may suggest sex-associated differences in comorbidity patterns, the underlying mechanisms could not be evaluated in the present study. Cross-dataset consistency was limited and, therefore, these findings should be interpreted cautiously.

Overall, while these EMR-derived comorbidity profiles provided meaningful insights into patterns of co-occurring conditions and the heterogeneity of AD, we acknowledge that administrative codes alone cannot fully capture the biological complexity in AD. Because AD and comorbidity diagnoses were based on administrative codes without biomarker or imaging confirmation, the study population may have included patients with mixed dementia, non-Alzheimer dementias, or miscoded diagnoses. These sources of diagnostic uncertainty may have influenced the observed clustering patterns. Future studies should focus on examining these findings in longitudinal datasets, integrating biomarkers and genetic data to better characterize patterns, and incorporating treatment-related variables such as medication use and treatment response. Additionally, psychosocial and lifestyle factors may also contribute to the observed clustering patterns and warrant investigation ^36–38^. Together, these efforts will be important to further assess the clinical relevance of comorbidity patterns and their role in understanding heterogeneity within the AD population.

### Final thoughts

Through unsupervised learning, we identified five dataset-derived comorbidity clusters and observed that AD-related comorbidities can be categorized into heterogeneous patterns, including clusters characterized by cardiovascular, gastrointestinal, and frailty-related conditions. Although these comorbidities correspond to well-recognized clinical syndromes, their enrichment within specific clusters indicates that they are not uniformly distributed among AD patients and may reflect heterogeneous comorbidity distributions across the AD population.

Future studies should investigate the factors underlying the observed comorbidity patterns and assess their reproducibility in longitudinal datasets. Incorporating longitudinal, biomarker, genomic and other biological data into such studies may help clarify the factors underlying the observed patterns.

Our findings may suggest that comorbidity patterns are relevant for understanding patient heterogeneity. Differences in comorbidity profiles may reflect variation in clinical care needs. However, their implications for treatment strategies remain unclear. These findings may contribute to a better understanding of heterogeneity within the AD population but should be interpreted as descriptive and hypothesis-generating.

## Limitations of the study

There are several limitations in this study. First, the presence of imbalanced data resulted in some odds ratios being disproportionately inflated. Extremely rare events may represent random variation or coding anomalies rather than true biological differences or clinically meaningful associations.

Second, cross-dataset consistency of some clusters was limited. This discrepancy may partly result from demographic differences or coding practices between the UCSF and UC-Wide cohorts, which could have influenced the distribution of AD severity and comorbidity patterns. Future multi-institutional studies with more diverse cohorts will be essential to further examine and refine these clusters. In addition, we did not apply cluster definitions derived from the UCSF dataset to directly classify patients in the UC-Wide cohort. This may limit the ability to define stable data-driven groupings. Furthermore, variability in data completeness and fragmentation of EHR data, particularly in resource-limited settings, may further affect the robustness of clustering results.

Third, the use of EMRs introduces several limitations. EMRs are subject to potential misclassification, missing data, and variability in coding practices. AD cases were identified based on diagnosis codes without biomarker, imaging, or neuropathological confirmation. Therefore, the study population may have included individuals with mixed dementia, non-Alzheimer dementias, or miscoded diagnoses, introducing the possibility of diagnostic misclassification. In addition, we excluded ICD-10-CM code F00.1 from the AD cohort to improve specificity. We also excluded patients with AD who had no recorded comorbidities beyond AD to ensure clustering interpretability and to focus on comorbidity patterns. In the UCSF cohort, 441 patients (5.0% of otherwise eligible AD cases) were excluded for this reason. These approaches may potentially limit generalizability and introduce selection bias toward patients with higher medical complexity. Due to the deidentification process, age data were truncated in both datasets. In the UCSF dataset, birth dates prior to 1930 were recoded to 1930, whereas individuals aged 91 years or older were assigned an age of 91 years in the UC-Wide dataset. Consequently, small age differences among older patient groups should be interpreted cautiously and descriptively. Furthermore, our analysis was limited to medical institutions in California, necessitating caution when extrapolating these findings to other regions or populations. Aggregated diagnoses in a single binary patient-level profile reflect cumulative coding patterns. This approach may limit interpretations of the findings in terms of disease progression or sequencing. Together with the cross-sectional design, this limits the ability to assess temporal relationships, disease progression, or causal inference. Future longitudinal studies are warranted to explore the timing of comorbidity onset relative to AD diagnosis and to better characterize the longitudinal stability of the observed comorbidity patterns.

Finally, while our study primarily focused on LOAD, the lack of EOAD analysis constrains the understanding of AD heterogeneity. Future research should extend the investigation to include EOAD populations to provide a more comprehensive understanding of AD cluster-derived patterns.

Overall, given the observational design, limited reproducibility, and the absence of genetic and microbiome data, these findings should be interpreted as descriptive and hypothesis-generating rather than definitive evidence of distinct biological subtypes. Despite these limitations, clustering of routinely collected EHR data may offer an accessible framework for characterizing comorbidity patterns and generating clinical hypotheses, particularly in resource-limited settings where more detailed biomarker data may not be available.

## Resource availability

### Lead contact

Requests for further information and resources should be directed to and will be fulfilled by the lead contact, Marina Sirota.

### Materials availability

This study did not generate new unique reagents.

### Data and code availability

The data reported in this study cannot be deposited in a public repository because they contain sensitive, de-identified patient information derived from UCSF EMR and UC-Wide databases. Access is restricted to UCSF-affiliated individuals and approved collaborators. To request access, individuals not affiliated with UCSF may establish an official collaboration with a UCSF-affiliated investigator by contacting the principal investigator, Marina Sirota. Requests for collaboration are typically processed within a few weeks. UCSF-affiliated individuals seeking access to the UCSF EMR database may contact UCSF’s Clinical and Translational Science Institute or the UCSF Information Commons team for more details. The UCDDP database is accessible only to UC researchers who have completed analyses within their respective UC institutions and have provided a justified rationale for scaling their analyses across multiple UC health centers. In addition, censored source data underlying cluster-specific comorbidity pair prevalence and cluster-specific comorbidity enrichment analyses are provided in Data S1 and are publicly available as of the date of publication.

All original code has been deposited at https://github.com/yukari-katsuhara/AD_Comorbidity_Patterns The GitHub repository contains scripts for all major analyses and figures, as well as software versions, analysis parameters, and documentation required for reproducibility.

Any additional information required to reanalyze the data reported in this paper is available from the lead contact upon request.

## Acknowledgements

This research was primarily supported by grants from the National Institute on Aging (NIA) (R01AG060393 and R01AG057683). The authors sincerely thank John Kornak, Thomas Hoffmann, Christopher Seaman, and all members of the Sirota Lab for their insightful discussions and valuable guidance throughout the preparation of this manuscript. The authors also thank the Center for Data-driven Insights and Innovation at UC Health (CDI2; https://www.ucop.edu/uc-health/functions/center-for-data-driven-insights-and-innovations-cdi2.html), for its analytical and technical support related to use of the UC Health Data Warehouse and related data assets, including the UC Data Discovery Platform (UCDDP).

## Author contributions

Conceptualization: Y.K., U.K., T.T.O., A.S.T., and M.S.

Methodology: Y.K., U.K., T.T.O., A.S.T., and M.S.

Data Curation: Y.K., U.K., T.T.O., A.S.T., and M.S.

Formal Analysis: Y.K.

Visualization: Y.K.

Writing – Original Draft: Y.K.

Writing – Review & Editing: Y.K., U.K., T.T.O., A.S.T., and M.S.

Supervision: A.S.T.

Funding Acquisition: M.S.

## Declaration of generative AI and AI-assisted technologies

During the preparation of this work, the authors used ChatGPT-4o (released on May 13, 2024, by OpenAI) in order to refine language and enhance readability. After using this tool or service, the authors reviewed and edited the content as needed and take full responsibility for the content of the publication.

## Supplemental information

Document S1. Figures S1–S5 and Tables S1–S4

Data S1. Excel file containing additional data related to Figures 3 and 6.

## STAR Methods

### KEY RESOURCES TABLE

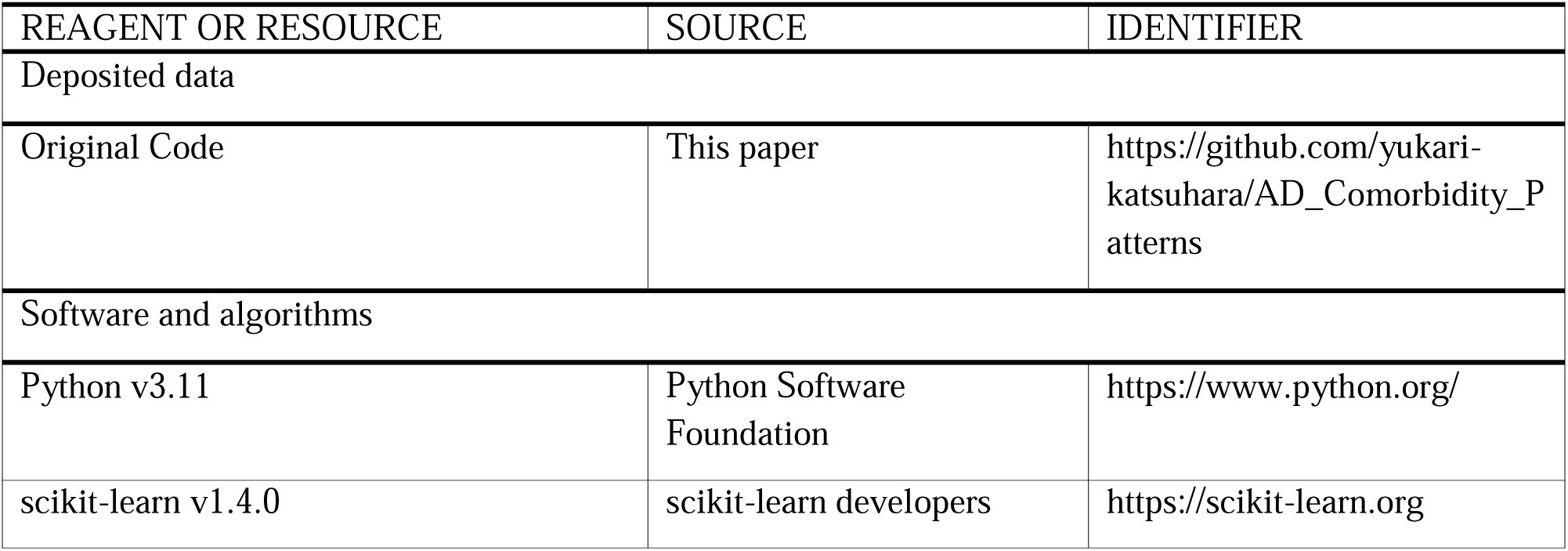
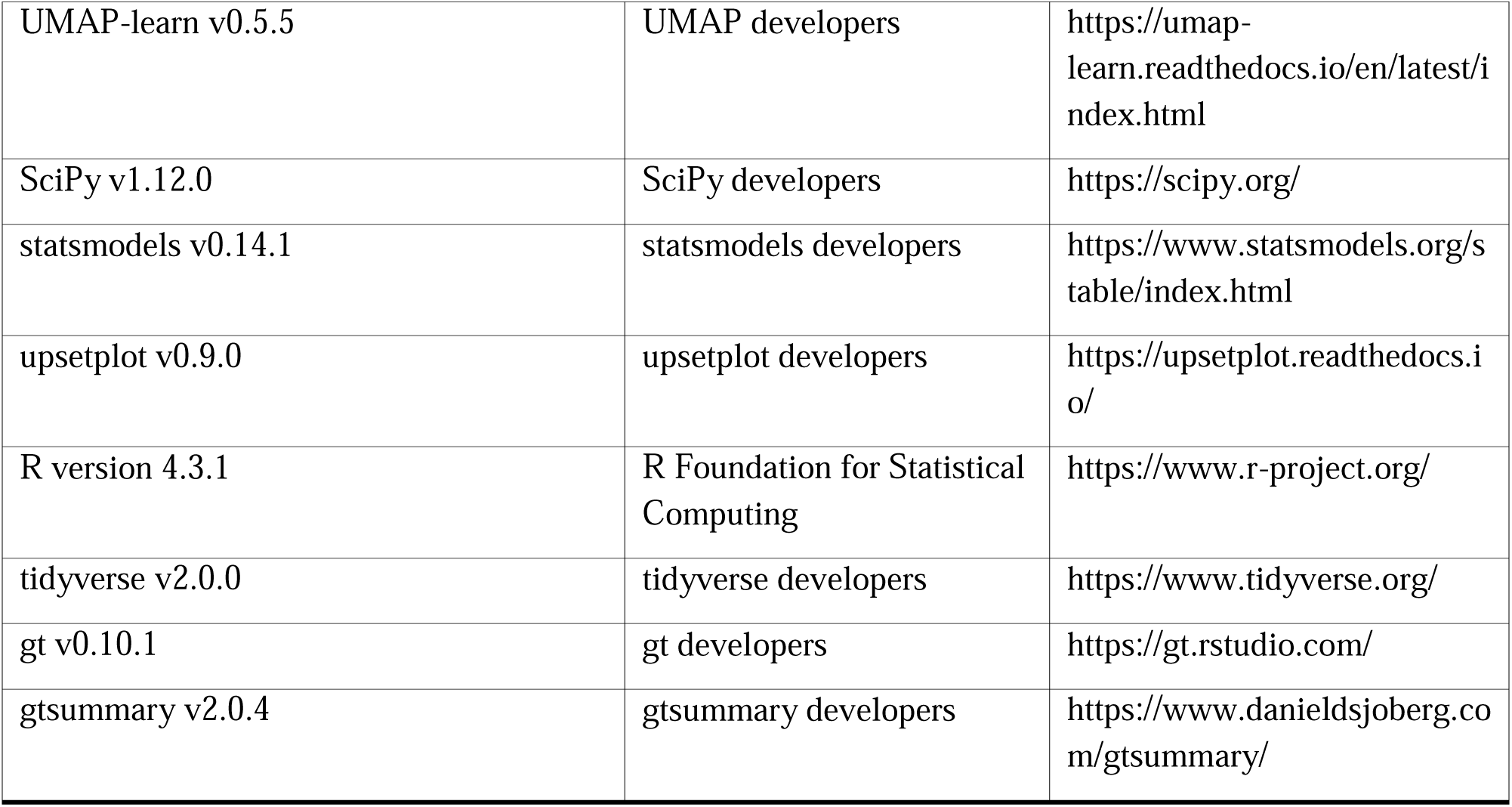

### EXPERIMENTAL MODEL AND STUDY PARTICIPANT DETAILS

#### Cohort identification

This is a retrospective cross-sectional study using EMR data. All clinical data used in this study were obtained from the University of California’s Health Data Warehouse (UCHDW) and the University of California Data Discovery Platform (UCDDP). UCDDP includes the de-identified records from UCSF, UC Los Angeles (UCLA), UC San Diego (UCSD), UC Davis (UCD), UC Irvine (UCI), and UC Riverside (UCR). This study identified comorbidity patterns of AD patients from UCSF EMR and evaluated their reproducibility using independent UC-Wide data from UCDDP.

Analyses of de-identified EMR data from UCSF and the UCDDP were conducted under Institutional Review Board (IRB) oversight from UCSF, UCD, UCI, UCLA, and UCSD. The UCSF IRB determined that this study did not meet the definition of human subjects research because only fully de-identified data were analyzed, and therefore IRB approval and the requirement for written informed consent were waived. This determination applied to the multi-institutional analysis using de-identified UCSF and UC-Wide datasets.

AD patients were identified from over five million records in the UCSF EMR dataset, which contains data from 1982 to 2020. Patients were included in the study if they met the inclusion criteria of an estimated age above 64 years and a diagnosis of AD based on the presence of the ICD-10-CM codes G30.1, G30.8, or G30.9. We focused on LOAD because it is a highly polygenic disorder, whereas EOAD is thought to have a distinct etiology and a stronger genetic predisposition ^39^. F00.1 was intentionally excluded because, in our dataset, it was frequently applied in the context of psychiatric or behavioral diagnoses and often encompassed vascular or mixed dementia cases. Including F00.1 could therefore introduce heterogeneity from non-AD etiologies, and its exclusion allowed us to define a more clinically homogeneous AD cohort, although this approach may potentially limit generalizability. Additionally, diagnosis records explicitly containing the term “Alzheimer” in the diagnosis name were excluded from the comorbidity feature set. Only patients with at least one remaining documented comorbidity in addition to AD were included in the analysis. This decision was made to ensure the stability of the clustering algorithm, as patients without documented comorbidities would be represented by all-zero vectors in the binary feature matrix. Moreover, because the primary objective of this study was to characterize comorbidity patterns, including patients without any comorbidities could dilute the signal within clusters and hinder the detection of meaningful patterns. Among 8,804 patients who otherwise met the AD eligibility criteria, 441 patients (5.0%) had no recorded comorbidity beyond AD and were excluded, resulting in a final analytic cohort of 8,363 AD patients.

This study was conducted as a cross-sectional analysis at the patient level. For each patient, diagnoses were aggregated across all available visits to create a single binary profile indicating the presence or absence of each diagnosis name. This approach ensured that each row in the dataset corresponded to an independent individual and mitigated the issue of correlated repeated measures across visits. However, this aggregation does not capture temporal changes in diagnoses and therefore does not allow assessment of time-specific or disease-stage-specific patterns. Sex was determined based on the most recent sex assignment recorded in the electronic medical record. The independent cohort was identified using the same criteria from UCDDP, excluding UCSF records, and consisted of UC-Wide clinical data from 2012 to 2024.

Due to de-identification procedures, age information was capped in both datasets. In the UCSF dataset, birth dates prior to 1930 were recoded to 1930. In the UC-Wide dataset, individuals aged 91 years or older were assigned an age of 91 years. Therefore, age-related analyses were limited to descriptive interpretation, and differences among the oldest age groups should be interpreted with caution.

Missing diagnoses were treated as absence of documentation and encoded as zeros in the binary diagnosis matrix. No imputation procedures were applied.

#### Sex as a biological variable

Our study examined both males and females using stratified analyses to identify sex differences in AD data-driven groupings. By conducting these analyses, we aimed to clarify potential sex-specific variations in disease characteristics.

### METHOD DETAILS

#### Dimensionality reduction and K-means clustering

After excluding diagnosis records explicitly containing the term “Alzheimer” in diagnosis names, one-hot encoding was performed for all diagnosis names. PCA was applied to diagnosis names to reduce the dimensionality of the dataset while preserving diagnostic information. PCA was implemented using scikit-learn with random_state=42. The number of retained components was 1000 for the UCSF dataset and 1100 for the UC-Wide dataset, corresponding to at least 80% cumulative explained variance. K-means clustering was then applied to these PCA components created from diagnosis names. K-means clustering was performed using the scikit-learn implementation with k-means++ initialization and random_state=42. All remaining parameters, including the number of initializations and convergence criteria, were set to the defaults of the scikit-learn version used for analysis. Cluster solutions were evaluated using silhouette scores, WSS reduction, the Calinski–Harabasz and Davies–Bouldin indices and interpretability considerations. While all clustering indices favored solutions with fewer clusters, the five-cluster solution was selected to provide additional granularity and improve interpretability of comorbidity patterns. Therefore, the five-cluster solution should be viewed as an interpretability-informed representation of the dataset rather than a uniquely optimal clustering solution. We also examined the top 10 pairs of co-occurring diagnoses ranked by their prevalence within each cluster to assess the clinical interpretability of different cluster configurations. To visualize the clustering results, patient distributions were projected onto a lower-dimensional space using UMAP.

#### Comorbidity enrichment analysis for overall trends of each cluster

To examine overall trends within each cluster, we performed enrichment analyses by comparing the frequency of each comorbidity in a given cluster with that in all other clusters combined. In this step, ICD-10-CM codes were used to define comorbidities, thereby facilitating clinical interpretation and improving the interpretability of cluster characteristics. For each diagnosis, the proportion of patients in the target cluster was compared to the proportion in the other clusters using Fisher’s exact test when the sample size was below five or the chi-squared test when the sample size was five or more. Statistical significance was determined using a Bonferroni-corrected threshold of p-value < 0.05. For each diagnosis, odds ratios, 95% confidence intervals, and p-values were calculated based on 2×2 contingency tables. The directionality of associations was determined based on the odds ratios. In addition to statistical significance, we calculated the prevalence of each comorbidity within each cluster to contextualize clinical relevance. Prevalence was defined as the proportion of patients within a given cluster with a specific diagnosis. Rare diagnoses were retained during clustering analyses. For interpretation of cluster characteristics, emphasis was placed on diagnoses with prevalence ≥5%, whereas lower-prevalence diagnoses were interpreted cautiously as exploratory signals. This allowed us to distinguish between statistically significant but low-frequency associations and more common comorbidities that may be more clinically meaningful. The statistical significance of associations and effect sizes of associations were visualized using volcano plots, while Manhattan plots (for the overall population) and Miami plots (for sex-stratified populations) illustrated the distribution of p-values across diagnoses. Sex-stratified analyses were performed to assess whether comorbidity patterns differed between male and female patients.

#### Comorbidity enrichment analysis for cluster- or cluster-sex-specific comorbidities

Cluster-specific comorbidities were identified by visualizing significant comorbidities using UpSet plots^40^. The statistical significance of these comorbidities was displayed in Manhattan plots for the overall population and Miami plots for sex-stratified populations. To further explore the phenotypic characteristics of each cluster, a UMAP visualization was generated using standardized odds ratios of cluster-specific comorbidities. Each dot represents a specific ICD-10-CM code, with its size corresponding to the odds ratio of the significantly associated cluster and its color indicating the respective cluster. The input features for the UMAP projection were derived from a matrix of standardized odds ratios in each cluster, where each row represents an ICD-10-CM code and each column corresponds to a cluster. Sex-stratified analyses were conducted to further investigate differences in comorbidity patterns between males and females.

#### Cross-dataset reproducibility assessment

To evaluate cross-dataset consistency of the clustering and comorbidity enrichment results, the same approach used for the UCSF dataset was applied to the UC-Wide dataset. AD patients were identified using the same inclusion criteria, and dimensionality reduction and K-means clustering were performed as described previously. Cluster-specific comorbidities in the UC-Wide dataset were analyzed using the same statistical methods and visualized with UpSet plots. The relationships between clusters in the UCSF and UC-Wide datasets were examined by comparing cluster-specific comorbidities, and the degree of overlap was assessed using Sankey plots. The coverage rate for UCSF clusters was calculated as the proportion of UCSF cluster-specific comorbidities that were statistically significant in UCSF and remained significant in UC-Wide clusters. The UC-Wide clusters with the highest coverage rates were identified as the best matches for each UCSF cluster. Log-log plots were used to compare the odds ratios of matched clusters between the UCSF and UC-Wide datasets for the overall population and sex-stratified subgroups. In addition, scatter plots were used to compare comorbidity prevalence between matched clusters. Pearson correlation analysis was performed to assess the linear relationship between the odds ratios, and the correlation coefficient and p-value were obtained to evaluate the statistical significance of the association.

### QUANTIFICATION AND STATISTICAL ANALYSIS

All statistical and computational analyses were conducted using Python, except for demographic table generation, which was performed in R. In the demographic table analysis, categorical variables such as sex, race, and death status were compared across clusters using the Chi-squared test, while continuous variables such as age and the number of comorbidities were analyzed using the Kruskal-Wallis rank sum test. When the Kruskal-Wallis test indicated a significant difference among clusters, post-hoc pairwise comparisons were conducted using Dunn’s test with Bonferroni correction to identify which specific groups differed significantly.

For comorbidity enrichment analysis, the proportions of patients with each ICD-10-CM code were compared using either Fisher’s exact test when fewer than five patients had the diagnosis and the chi-squared test when five or more patients had the diagnosis. Odds ratios and their 95% confidence intervals were calculated to quantify the strength and direction of associations. Diagnoses were considered significant if the p-value was below 0.05 after Bonferroni correction, and the directionality of the association was determined using the odds ratio.

