## Supplemental Figures for "Comorbidity patterns and sex differences in late-onset Alzheimer’s disease using electronic records"

**Supplemental Materials**

|  | UCSF | | | | | | | | |
| --- | --- | --- | --- | --- | --- | --- | --- | --- | --- |
| Number of Cluster | WSS | ΔWSS (%) | Silhouette | ΔSilhouette (%) | Calinski Harabasz | ΔCH (%) | Davies Bouldin | ΔDB (%) | Smallest Cluster (%) |
| 2 | 319553 | – | 0.439 | - | 497.64 | – | 3.7447 | – | 17.0%  (n=1424) |
| 3 | 312922 | 2.1% | 0.284 | 35.3% | 342.64 | 31.2% | 4.6098 | 23.1% | 8.8%(n=735) |
| 4 | 308949 | 1.3% | 0.272 | 4.2% | 267.16 | 22.0% | 4.8371 | 4.9% | 4.8%(n=403) |
| 5 | 306584 | 0.8% | 0.159 | 41.5% | 218.01 | 18.4% | 5.1402 | 6.3% | 3.9%(n=328) |
| 6 | 305359 | 0.4% | 0.019 | 88.1% | 181.8 | 16.6% | 5.6721 | 10.4% | 3.9%(n=322) |
|  | UC-Wide | | | | | | | | |
| Number of Cluster | WSS | ΔWSS (%) | Silhouette | ΔSilhouette (%) | Calinski Harabasz | ΔCH (%) | Davies Bouldin | ΔDB (%) | Smallest Cluster (%) |
| 2 | 1108599 | - | 0.273 | - | 1774.358 | – | 3.6563 | – | 27.0%  (n=6996) |
| 3 | 1089258 | 1.7% | 0.141 | 48.4% | 1132.773 | 36.2% | 5.1086 | 39.7% | 12.0%  (n=3111) |
| 4 | 1078519 | 1.0% | 0.136 | 3.5% | 848.604 | 25.1% | 5.4058 | 5.8% | 10.1%  (n=2620) |
| 5 | 1071218 | 0.7% | 0.068 | 50.0% | 683.732 | 19.4% | 6.0116 | 11.2% | 6.8%  (n=1765) |
| 6 | 1065004 | 0.6% | 0.048 | 29.4% | 581.301 | 15.0% | 5.9059 | -1.8% | 3.7%(n=949) |

**Supplemental Table 1. Cluster performance metrics across clustering solutions**

WSS, Within-cluster Sum of Squares; CH, Calinski Harabasz; DB, Davies Bouldin

| **4 Clusters** | | | | | | | | | | | | | | |
| --- | --- | --- | --- | --- | --- | --- | --- | --- | --- | --- | --- | --- | --- | --- |
| Cluster 1 (N=403) | | | Cluster 2 (N=1,293) | | | Cluster 3 (N=817) | | | Cluster 4 (N=5,850) | | |  |  |  |
| ICD-10-CM | | Prev. | ICD-10-CM | | Prev. | ICD-10-CM | | Prev. | ICD-10-CM | | Prev. |  |  |  |
| I10 | Z00.00 | 0.043 | I10 | Z78.9 | 0.098 | F02.80 | I10 | 0.068 | I10 | Z00.00 | 0.043 |  |  |  |
| I10 | Z23 | 0.043 | I10 | R07.9 | 0.093 | F02.80 | F03.90 | 0.066 | I10 | Z23 | 0.043 |  |  |  |
| Z00.00 | Z23 | 0.043 | I10 | N39.0 | 0.092 | F03.90 | I10 | 0.062 | Z00.00 | Z23 | 0.043 |  |  |  |
| E78.5 | I10 | 0.041 | D64.9 | I10 | 0.085 | E78.5 | F02.80 | 0.056 | E78.5 | I10 | 0.041 |  |  |  |
| E78.5 | Z00.00 | 0.039 | F06.8 | I10 | 0.084 | E78.5 | I10 | 0.053 | E78.5 | Z00.00 | 0.039 |  |  |  |
| E78.5 | Z23 | 0.039 | E78.5 | I10 | 0.082 | F02.80 | Z00.00 | 0.049 | E78.5 | Z23 | 0.039 |  |  |  |
| I10 | R05 | 0.039 | F03.90 | I10 | 0.080 | E78.5 | F03.90 | 0.048 | I10 | R05 | 0.039 |  |  |  |
| I10 | R07.9 | 0.038 | I10 | Z00.00 | 0.078 | F02.80 | Z71.89 | 0.048 | I10 | R07.9 | 0.038 |  |  |  |
| R05 | Z00.00 | 0.038 | I10 | Z23 | 0.073 | F02.80 | Z23 | 0.047 | R05 | Z00.00 | 0.038 |  |  |  |
| R05 | Z23 | 0.038 | R53.81 | R53.83 | 0.071 | Z00.00 | Z23 | 0.047 | R05 | Z23 | 0.038 |  |  |  |
| **5 Clusters** | | | | | | | | | | | | | | |
| Cluster 1 (N=328) | | | Cluster 2 (N=538) | | | Cluster 3 (N=815) | | | Cluster 4 (N=1,521) | | | Cluster 5 (N=5,161) | | |
| ICD-10-CM | | Prev. | ICD-10-CM | | Prev. | ICD-10-CM | | Prev. | ICD-10-CM | | Prev. | ICD-10-CM | | Prev. |
| I10 | Z00.00 | 0.035 | I10 | R07.9 | 0.053 | F02.80 | I10 | 0.068 | I10 | Z78.9 | 0.080 | F02.80 | I10 | 0.083 |
| Z00.00 | Z23 | 0.035 | E78.5 | I10 | 0.050 | F02.80 | F03.90 | 0.066 | I10 | N39.0 | 0.080 | F02.80 | F03.90 | 0.071 |
| I10 | Z23 | 0.035 | I10 | Z78.9 | 0.049 | F03.90 | I10 | 0.062 | F03.90 | I10 | 0.077 | F03.90 | I10 | 0.066 |
| E78.5 | I10 | 0.033 | I10 | Z00.00 | 0.048 | E78.5 | F02.80 | 0.056 | F06.8 | I10 | 0.068 | E78.5 | I10 | 0.062 |
| E78.5 | Z00.00 | 0.032 | I10 | N39.0 | 0.045 | E78.5 | I10 | 0.054 | D64.9 | I10 | 0.065 | E78.5 | F02.80 | 0.055 |
| E78.5 | Z23 | 0.032 | D64.9 | I10 | 0.045 | F02.80 | Z00.00 | 0.050 | I10 | R07.9 | 0.063 | I10 | R41.3 | 0.049 |
| I10 | R05 | 0.031 | I10 | Z23 | 0.045 | E78.5 | F03.90 | 0.048 | E78.5 | I10 | 0.058 | F02.80 | R41.3 | 0.047 |
| R05 | Z00.00 | 0.031 | R53.81 | R53.83 | 0.043 | F02.80 | Z23 | 0.048 | F03.90 | N39.0 | 0.055 | F02.80 | G31.84 | 0.045 |
| F02.80 | I10 | 0.031 | I10 | R53.81 | 0.043 | Z00.00 | Z23 | 0.048 | F03.90 | Z78.9 | 0.051 | F02.80 | Z66 | 0.042 |

**Supplemental Table 2. Cluster-specific comorbidity pair prevalence for K = 4 and K = 5**

Prev., Prevalence

ICD-10-CM, the International Classification of Diseases, 10th Revision, Clinical Modification.

| Age in UCSF Cohort | | | | | Age in UC-Wide Cohort | | | | |
| --- | --- | --- | --- | --- | --- | --- | --- | --- | --- |
|  | Cluster 5 | Cluster 1 | Cluster 2 | Cluster 3 |  | Cluster A | Cluster B | Cluster C | Cluster D |
| Cluster 1 | 0.80 | - | - | - | Cluster B | 1 | - | - | - |
| Cluster 2 | 0.00* | 0.00* | - | - | Cluster C | 0.00* | 0.00* | - | - |
| Cluster 3 | 0.00* | 0.00* | 0.00* | - | Cluster D | 0.00* | 0.00* | 0.00* | - |
| Cluster 4 | 0.00* | 0.00* | 0.38 | 0.00* | Cluster E | 0.73 | 0.45 | 0.00* | 0.00* |
| Number of comorbidities in UCSF Cohort | | | | | Number of comorbidities in UC-Wide Cohort | | | | |
|  | Cluster 5 | Cluster 1 | Cluster 2 | Cluster 3 |  | Cluster A | Cluster B | Cluster C | Cluster D |
| Cluster 1 | 0.00* | - | - | - | Cluster B | 0.00* | - | - | - |
| Cluster 2 | 0.00* | 0.00* | - | - | Cluster C | 0.00* | 0.00* | - | - |
| Cluster 3 | 0.00* | 0.00* | 0.10 | - | Cluster D | 0.00* | 0.00* | 0.00* | - |
| Cluster 4 | 0.00* | 0.00* | 0.00* | 0.00* | Cluster E | 0.00* | 0.00* | 0.00* | 0.00* |
| Sex in UCSF Cohort | | | | | Sex in UC-Wide Cohort | | | | |
|  | Cluster 5 | Cluster 1 | Cluster 2 | Cluster 3 |  | Cluster A | Cluster B | Cluster C | Cluster D |
| Cluster 1 | 0.67 | - | - | - | Cluster B | 0.00* | - | - | - |
| Cluster 2 | 0.16 | 1 | - | - | Cluster C | 0.00* | 0.00* | - | - |
| Cluster 3 | 1 | 1 | 1 | - | Cluster D | 0.00* | 0.00* | 1 | - |
| Cluster 4 | 0.00* | 0.94 | 0.49 | 0.00* | Cluster E | 0.19 | 0.00* | 0.00* | 0.00* |

**Supplemental Table 3. Bonferroni-corrected p-values for pairwise comparisons across clusters**

P-values for age and the number of comorbidities were calculated using the Kruskal-Wallis rank sum test with Bonferroni correction. P-values for pairwise comparisons of sex were obtained from proportion tests with Bonferroni correction. *, Adjusted p-value < 0.05; UC, University of California

|  | **Cluster 1, *N* = 328**  **(Sex: Unknown N=1)** | | | | **Cluster 2, *N* = 538**  **(Sex: Unknown N=0)** | | | | **Cluster 3, *N* = 815**  **(Sex: Unknown N=0)** | | | |
| --- | --- | --- | --- | --- | --- | --- | --- | --- | --- | --- | --- | --- |
| Sex, N, *n* (%) | Female  (N=217) | | Male  (N=110) | | Female (N=358) | | Male  (N=180) | | Female (N=514) | | Male  (N=301) | |
| Race, *n* (%) |  |  |  |  |  |  |  |  |  |  |  |  |
| White or Caucasian | 91 | (42%) | 57 | (52%) | 150 | (42%) | 98 | (54%) | 268 | (52%) | 171 | (57%) |
| Other/Unknown | 31 | (14%) | 13 | (12%) | 68 | (19%) | 30 | (17%) | 46 | (8.9%) | 29 | (9.6%) |
| Asian | 71 | (33%) | 37 | (34%) | 65 | (18%) | 25 | (14%) | 143 | (28%) | 79 | (26%) |
| Black or African American | 24 | (11%) | 3 | (2.7%) | 53 | (15%) | 17 | (9.4%) | 47 | (9.1%) | 18 | (6.0%) |
| Native Hawaiian or Other Pacific Islander | 0 | 0 |  |  | 22 | (6.1%) | 10 | (5.6%) | 10 | (1.9%) | 4 | (1.3%) |
| Death Status, *n* (%) |  |  |  |  |  |  |  |  |  |  |  |  |
| Alive | 145 | (67%) | 73 | (66%) | 176 | (49%) | 82 | (46%) | 358 | (70%) | 192 | (64%) |
| Deceased | 72 | (33%) | 37 | (34%) | 182 | (51%) | 98 | (54%) | 156 | (30%) | 109 | (36%) |
| Age (yr), Median (IQR) | 90 | (84-91) | 89 | (84-91) | 90 | (90-91 | 90 | (90-91) | 88 | (82.3-90) | 87 | (81-90) |
| Comorbidities, Median (IQR) | 313 | (256-373) | 314 | (265.3-388.8) | 129 | (105-168) | 138.5 | (108.5-183) | 112 | (78.3-156.8) | 118 | (84-155) |

|  | **Cluster 4, *N* = 1,521**  **(Sex: Unknown N=0)** | | | | **Cluster 5, *N* = 5,161**  **(Sex: Unknown N=18)** | | | |
| --- | --- | --- | --- | --- | --- | --- | --- | --- |
| Sex, *n* (%) | Female (N=1,083) | | Male  (N=438) | | Female (N=3,143) | | Male  (N=2,000) | |
| Race, *n* (%) |  |  |  |  |  |  |  |  |
| White or Caucasian | 632 | (58%) | 265 | (61%) | 3,171 | (60%) | 1,400 | (70%) |
| Other/Unknown | 142 | (13%) | 63 | (14%) | 849 | (16%) | 307 | (15%) |
| Asian | 74 | (6.8%) | 35 | (8.0%) | 582 | (11%) | 118 | (5.9%) |
| Black or African American | 148 | (14%) | 32 | (7.3%) | 431 | (8.1%) | 81 | (4.1%) |
| Native Hawaiian or Other Pacific Islander | 87 | (8.0%) | 43 | (9.8%) | 282 | (5.3%) | 94 | (4.7%) |
| Death Status, *n* (%) |  |  |  |  |  |  |  |  |
| Alive | 635 | (59%) | 277 | (63%) | 3,959 | (74%) | 1,681 | (84%) |
| Deceased | 448 | (41%) | 161 | (37%) | 1,356 | (26%) | 319 | (16%) |
| Age (yr), Median (IQR) | 91 | (90-91) | 90 | (90-91) | 90 | (82-91) | 88 | (80-91) |
| Comorbidities, Median (IQR) | 46 | (32-64) | 49 | (35-66.8) | 12 | (5-21) | 12 | (5-22) |

**Supplemental Table 4. Sex-stratified patient demographics in UCSF.** Medians and interquartile ranges are presented as median (25th percentile-75th percentile). "Comorbidities" refers to "Number of comorbidities," which represents the total count of diagnoses per patient. The category "Unknown" in Sex includes records classified as “Unknown.” The category "Other/Unknown" in Race includes records classified as “American Indian or Alaska Native,” “Other,” “0,” “Unknown/Declined,” “Unknown,” and “Declined.”

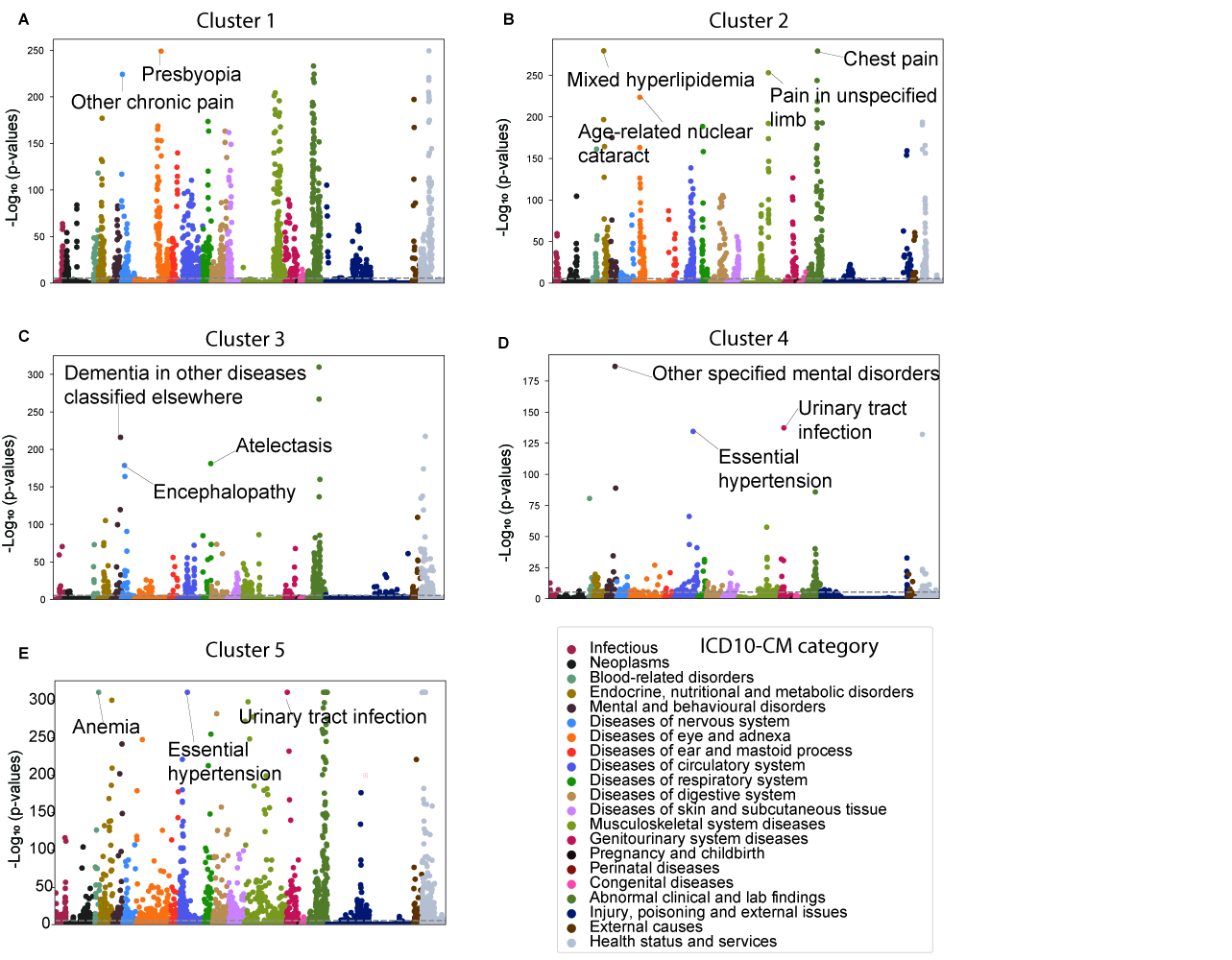

**Supplemental Figure 1. Manhattan plots of cluster-specific comorbidity enrichment patterns.** Manhattan plots displaying enriched ICD-10-CM codes identified using the two-sided Fisher’s exact test (for cases < 5) or the Chi-squared test (for cases ≥ 5). Enrichment was determined based on a Bonferroni-corrected p-value < 0.05. ICD-10-CM, the International Classification of Diseases, 10th Revision, Clinical Modification. **A:** Cluster 1, **B:** Cluster 2, **C:** Cluster 3, **D:** Cluster 4, **E:** Cluster 5

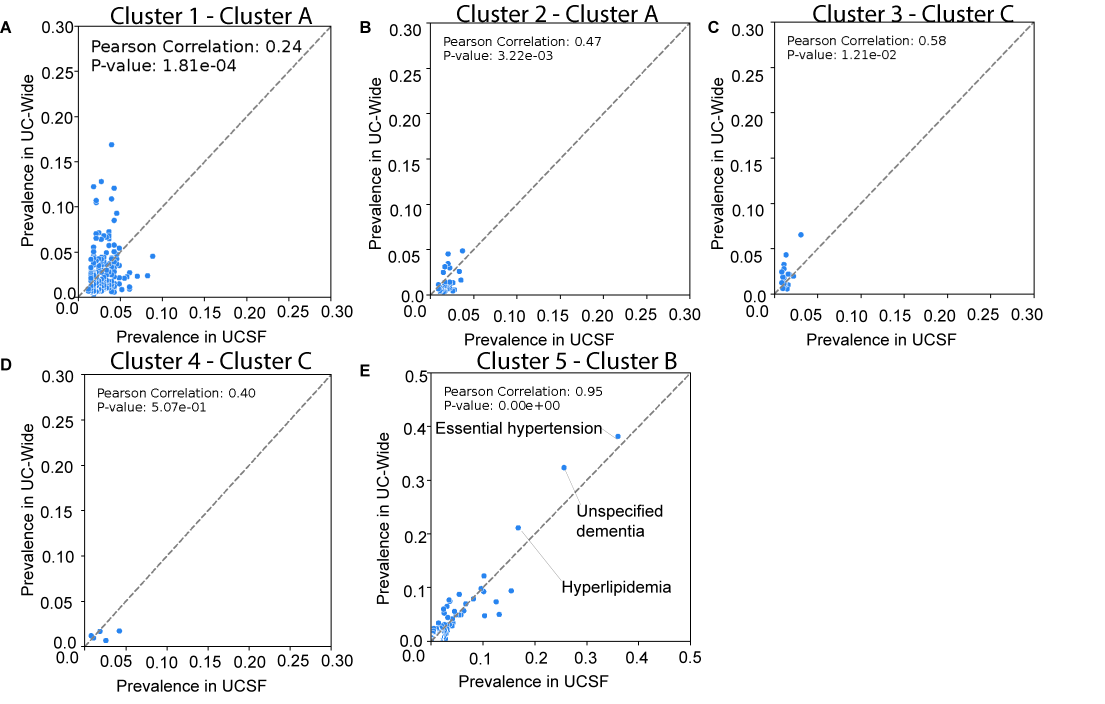

**Supplemental Figure 2. Cross-dataset comparison of comorbidity prevalence.** Comparison of comorbidity prevalence showing partial reproducibility of comorbidity patterns across the UCSF and UC-Wide datasets, particularly in Cluster 5.

**A:** Cluster 1 vs. Matched UC-Wide Cluster A, **B:** Cluster 2 vs. Matched UC-Wide Cluster A, **C:** Cluster 3 vs. Matched UC-Wide Cluster C, **D:** Cluster 4 vs. Matched UC-Wide Cluster C, **E:** Cluster 5 vs. Matched UC-Wide Cluster B

P-values were calculated from Pearson correlation analysis. UC, University of California.

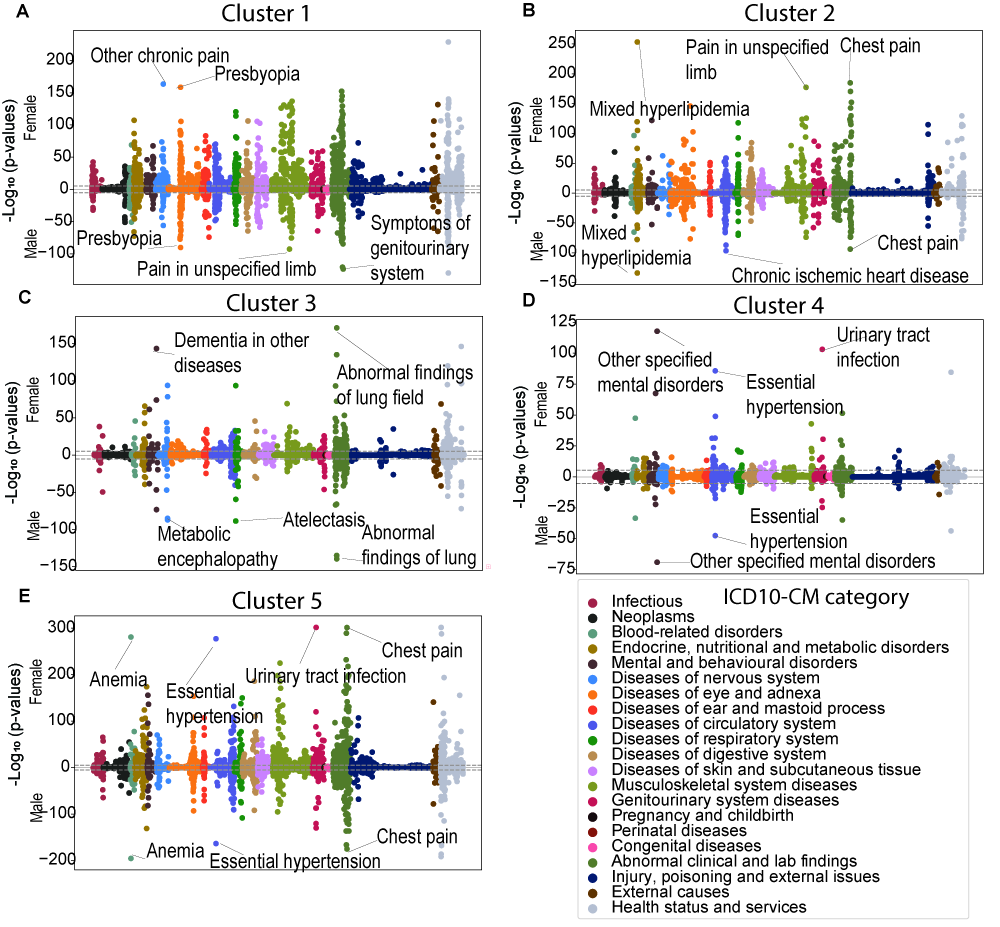

**Supplemental Figure 3. Miami plots of sex-stratified comorbidity enrichment patterns across clusters.**

Miami plots displaying enriched ICD-10-CM codes identified using the two-sided Fisher’s exact test (for cases < 5) or the Chi-squared test (for cases ≥ 5). Enrichment was determined based on a Bonferroni-corrected p-value < 0.05. ICD-10-CM, the International Classification of Diseases, 10th Revision, Clinical Modification. **A:** Cluster 1, **B:** Cluster 2, **C:** Cluster 3, **D:** Cluster 4, **E:** Cluster 5

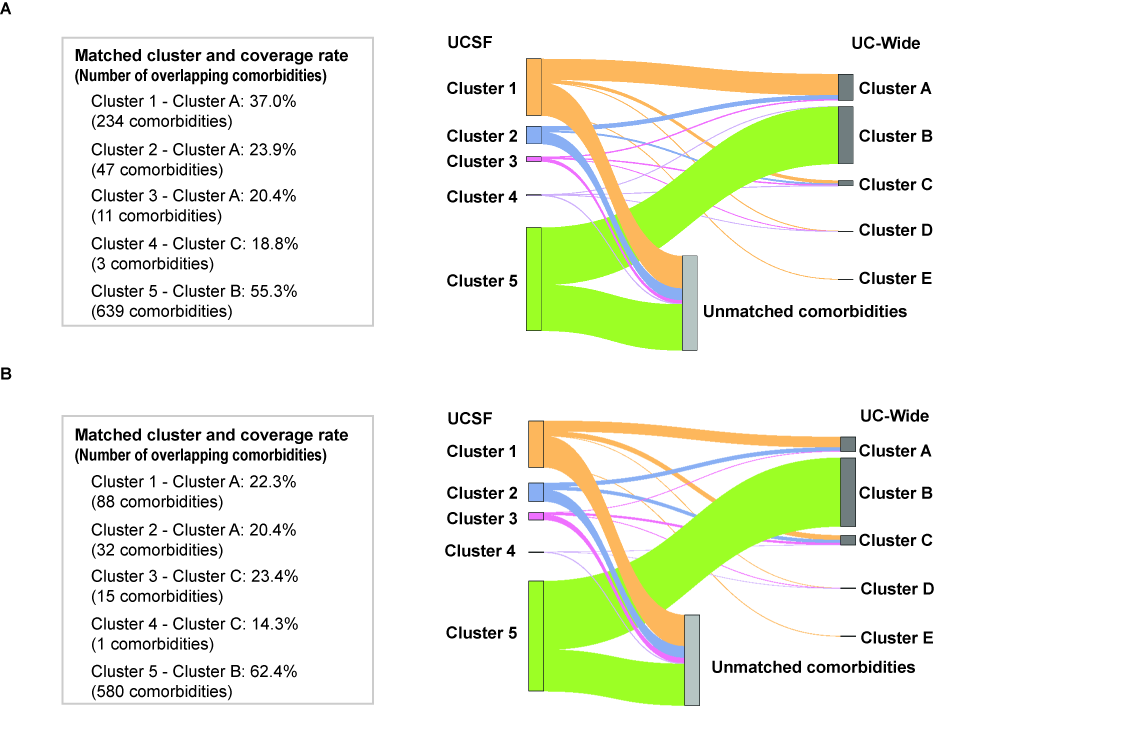

**Supplemental Figure 4. Sex-stratified Sankey diagram and coverage rate**

Sankey plot illustrating the partial overlap of significant comorbidities across UCSF and UC-Wide populations within each cluster. The coverage rate represents the proportion of UCSF cluster-specific comorbidities that remain significant in both the UCSF and UC-Wide datasets, identifying the most closely matched UC cluster. UC, University of California.

**A:** Female. Number of cluster-specific comorbidities: Cluster 1 (633), Cluster 2 (197), Cluster 3 (54), Cluster 4 (16), Cluster 5 (1,156), Cluster A (1,776), Cluster B (2,108), Cluster C (576), Cluster D (24), Cluster E (10)

**B:** Male. Number of cluster-specific comorbidities: Cluster 1 (395), Cluster 2 (157), Cluster 3 (64), Cluster 4 (7), Cluster 5 (929), Cluster A (1,053), Cluster B (1,660), Cluster C (604), Cluster D (33), Cluster E (13)

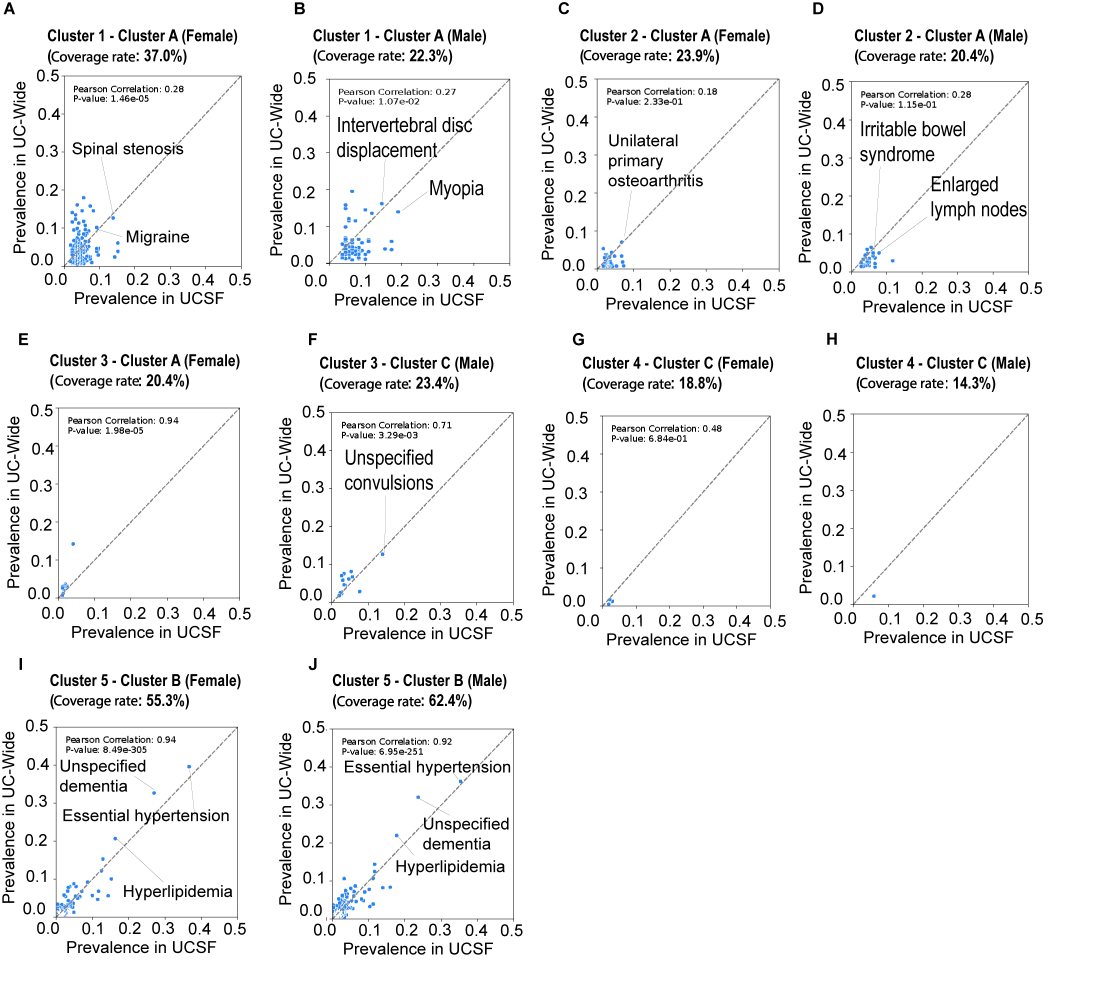

**Supplemental Figure 5. Sex-stratified cross-dataset comparison of comorbidity prevalence.**

Comparison of comorbidity prevalence showing partial reproducibility of comorbidity patterns across the UCSF and UC-Wide datasets, particularly for Clusters 1 and 5.

The covered rate represents the proportion of UCSF cluster-sex-specific comorbidities that remain significant in both the UCSF and UC-Wide datasets. **A, C, E, G, I**: Cluster 1–Cluster 5 (Female) vs. Matched UC-Wide Cluster (Female), **B, D, F, H, J:** Cluster 1–Cluster 5 (Male) vs. Matched UC-Wide Cluster (Male). P-values were calculated from Pearson correlation analysis. UC, University of California.
